# Diagnostic accuracy and testing efficiency of pooled testing of sputum swabs to detect tuberculosis on the near point of care Pluslife MiniDock MTB assay

**DOI:** 10.64898/2026.08.05.26359243

**Authors:** Nadege Bonkar Chifu, Asonganyi Etiendem, Diana Kolieghu Tcheumeni, Angela Neh, Ngha Ndze Mbuh, Gilford Fonyuy, Denis Nsame, Norah Nyah Ndi, Irene Adeline Goupeyou Wandji, Mercy Fundoh, Cyrille Mbuli, Nestor Biatu, Comfort Vuchas, Tushar Garg, Jacob Creswell, Melissa Sander, the RAPID TB team

## Abstract

**Background:** Pooled testing increases testing efficiency and reduces testing costs. This approach has been recently recommended by the World Health Organization for use with low-complexity nucleic acid amplification TB diagnostics to increase access to testing when resources are constrained. Pooled testing can also be used with novel near point of care tests, and evidence is needed on diagnostic performance of pooled testing in these more portable, lower cost tests.

**Methods:** We evaluated pooled testing on the Pluslife MiniDock MTB assay with stored sputum collected from adults with presumptive TB. We assessed sensitivity and specificity against the reference standard of liquid culture and diagnostic agreement against Xpert MTB/RIF Ultra and individual MiniDock MTB; we also estimated pooled testing efficiency.

**Results:** Swabs from sputum specimens were tested in 287 pools of 3 and on 861 individual tests. Against culture, sensitivity of testing was 88% (87/99, 95%CI, 80-93%) as compared to 89% (88/99, 95%CI, 81-94%) for individual MiniDock MTB testing, with pooled testing specificity of 99% (97-99%) as compared to 95% (94-97%) for individual testing. Pooled testing saved 32% of tests in this population that included 12% (100) people with culture-positive TB.

**Conclusions:** Pooled testing with sputum swabs from three people had similar diagnostic accuracy against TB culture as individual sputum swab testing in this evaluation. These results provide evidence that pooled testing with near point of care tests could help to further reduce testing costs and help to expand access to molecular testing at the lowest levels of the health system.

## INTRODUCTION

In early 2026, the World Health Organization (WHO) released new recommendations for TB diagnostics, including recommending near point of care nucleic acid amplification test (NPOC-NAAT) testing with the MiniDock MTB assay (Guangzhou Pluslife Biotech, China) to replace smear microscopy, and recommending the use of pooled testing for tuberculosis (TB) on the low-complexity automated NAAT (LC-aNAAT) GeneXpert (Cepheid Sunnyvale, USA) platform [1]. Both recommendations address the need to improve access to molecular testing for people with presumptive TB, as in 2024 only about half of people with TB received a molecular test at the time of diagnosis [2]. The Xpert MTB/RIF Ultra (Ultra) assay has higher sensitivity than the MiniDock MTB, but the Ultra assay also costs more than twice as much per test, and the testing equipment is much more costly [3–6]. A recent multicentre evaluation demonstrated that pooled testing of sputum samples on the Ultra could save more than 50% of test costs across a diverse population, with a drop in sensitivity of about 3% as compared to individual Ultra testing [7].

Pooled testing has been implemented in many disease areas and is often used to scale up testing when resources are constrained, as for COVID-19 [8–12], TB [13–16], and other infectious diseases [17–19], driven by realities in laboratories that face supply chain challenges and budget shortfalls.

Typically, pooled testing is performed by pooling a portion of each specimen together prior to testing, resulting in dilution of the target to be detected and leading to reduced sensitivity compared to individual testing. When pooled testing is conducted with swabs, one approach is to pool the material from the swabs at the elution stage, and in this case the specimen is not necessarily diluted. Pooled swab testing has been used for detection of influenza and SARS-CoV-2, with no dilution reported[20,21]. However, when using this approach, the amount of material that can be added may be limited, depending on the volume and type of specimen and the assay; also, when the swab is taken directly from the patient, it may be necessary to collect two swabs initially or to collect a second swab for follow-on testing if the pool is positive. For pooled testing with sputum swabs, no additional specimen collection is necessary, since swabs for individual follow-on testing after a positive pool can be prepared from the same initial sputum specimen.

While there are only a few published studies on the diagnostic performance of MiniDock MTB, there are not yet studies evaluating how pooled testing could be performed with the assay to increase testing efficiency and reduce testing cost and time. Here, we conducted a diagnostic accuracy study on the performance of pooled testing using MiniDock MTB to assess the potential of this approach.

## METHODS

### Study design and population

This was a diagnostic accuracy study to assess pooled testing on the Pluslife MiniDock MTB assay using stored sputum specimens collected from participants with presumptive TB who were enrolled in a multisite study that was carried out at five health facilities and communities in Cameroon from February to June 2025 (see Supplemental Methods) [4].

Retrospective testing on stored sputum specimens was conducted at the Tuberculosis Reference Laboratory Bamenda, which is accredited in accordance with the recognized International Standard ISO 15189:2022 (SANAS Accredited Medical Laboratory, no. M0593).

This secondary study was approved by the Institutional Review Board of the Cameroon Baptist Convention Health Board. The original study was approved by the National Ethics Committee for Research on Human Health and by the Institutional Review Board of the Cameroon Baptist Convention Health Board, with written informed consent provided by all participants. The Standards for Reporting of Diagnostic Accuracy (STARD) recommendations were followed for reporting these results [22]. Analyses were performed in R version 4.6.0.

## Procedures

### Pluslife MiniDock MTB testing-individual tests and pools of three

For this evaluation, a pool size of 3 was used (see Supplemental Methods for pool size determination). Frozen sputum aliquots (0.5mL) were first thawed to room temperature. Sputum swabs were prepared following the manufacturer’s instructions. Briefly, a swab was swirled in sputum ten times over 10-15 seconds, ensuring that the swab head reached each area of the sputum container. Then the swab was eluted into the Releasing Agent tube as shown in Figure 1.

**Figure 1.**
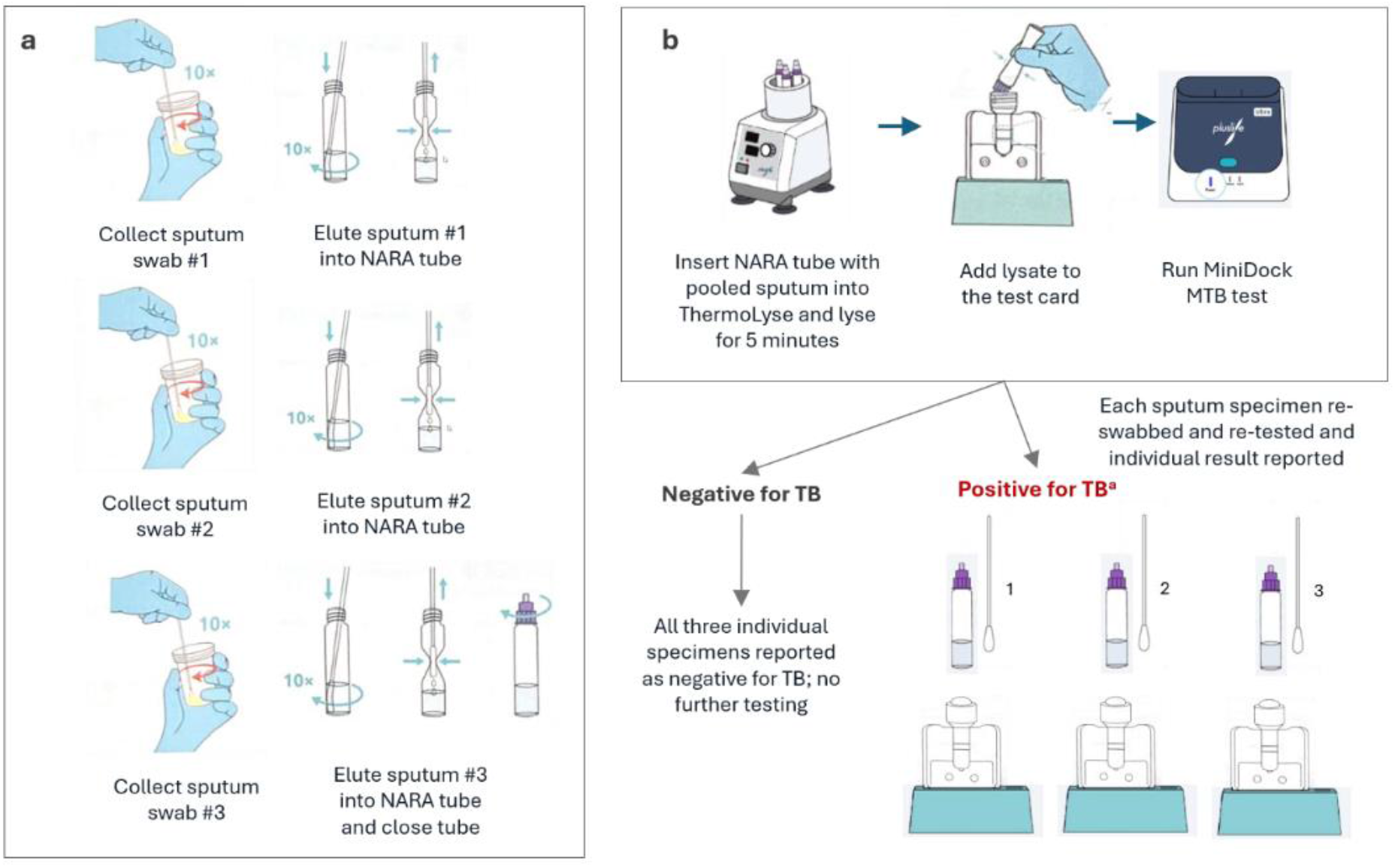
Schematic of pooled testing procedure for pools of three sputum swabs on the MiniDock MTB assay. (a) Three sputum swabs prepared from three different sputum specimens are pooled at the elution step. The first swab is prepared by swirling the swab in circles in the sputum at least 10 times for 10-15 seconds; then the swab with the sputum is inserted into the nucleic acid releasing agent (NARA) tube and the swab tip is twisted against the bottom and sides of the tube 10 times, while squeezing the tube wall to maximize elution of the sputum from the swab. Next the first sputum container is closed, and the second is opened, and the second swab is prepared and eluted the same way as the first; this is repeated for the third swab, and then the NARA tube is closed. (b) The NARA tube with the pooled sputum is added to the Pluslife Thermolyse and lysed for 5 minutes at 3000rpm and 75°C, following the manufacturer’s instructions. The lysate is then added to the MTB test card, allowed to stand for 15 seconds, then the airbag at the top of the card is pressed and the card is shaken up and down 10 times in 5 seconds. Then the card is inserted into the MiniDock and tested as usual. For pools with a result of negative, the result of each individual specimen is reported as negative, with no further testing. For pools with a result of positive (or invalid), another sputum swab from the same sputum specimen is prepared and tested individually on the MiniDock MTB, and this individual result is reported for the specimen.

**Figure 2.**
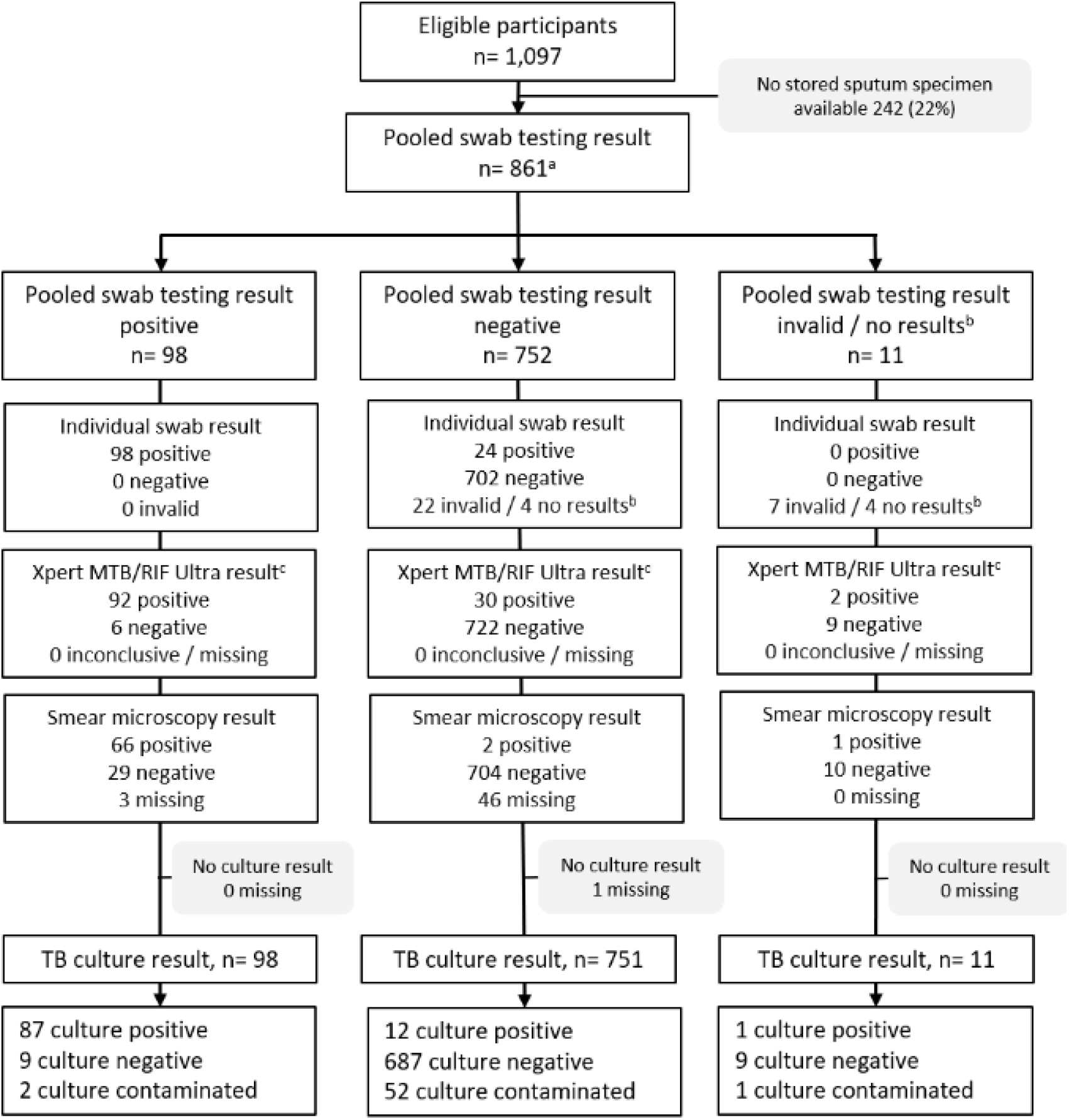
STARD flow. ^a^Specimens from 6 people among the total 855 participants were tested in two pools each; these specimens each had negative results on both individual results. ^b^8 specimens had no results on individual testing as the remaining sputum volume was deemed insufficient to perform individual testing. ^c^Ultra testing was performed on a different sputum specimen from the same participant (Figure S1); final Ultra result (after re-testing for invalid initial results).

Three sputum swabs prepared from three different specimens were pooled at the elution step. To create a pool, the three swabs were eluted in the same tube serially.

Two sputum swabs were prepared from each sputum specimen; one was tested individually and the other was used to create a pool with swabs from two other specimens.

After preparing the pool, tests were performed following manufacturer’s instructions (Supplemental Methods). The MiniDock MTB assay provides results of positive, negative or invalid.

Specimens with invalid results were not repeated. Pooled and individual MiniDock MTB tests were performed by technicians without knowledge of any other TB test results or clinical information.

## Data analysis

The pooled test result for each specimen was classified as negative if the pool result was negative; if the result of the pool was positive or invalid, then the individual test result was used as the pooled test result for the specimen (**Figure 1**).

For the diagnostic accuracy analysis, a microbiological reference standard (MRS) based on culture from one sputum specimen was used, including one automated liquid culture. Participants for whom at least one culture was positive for TB were defined as having culture-positive TB; among these, anyone with at least one smear positive for TB was defined as having smear positive TB. Participants for whom MGIT or both cultures were negative for TB were defined as culture negative for TB; participants for whom culture results were contaminated or missing were excluded from the analysis. For the Ultra comparator, any result with MTB detected was considered positive.

A diagnostic agreement analysis of the MiniDock MTB pooled test results was performed against the Ultra comparator and against the MiniDock MTB individual results.

Participants were characterized using simple descriptive statistics. To evaluate the diagnostic accuracy of the pooled MiniDock MTB against the MRS, we calculated the sensitivity and specificity point estimates with 95% confidence intervals (CIs) using the Wilson score method; the same approach was used to obtain positive and negative percent agreement estimates against the Ultra and individual MiniDock MTB comparator tests. The proportion of non-actionable results for the pooled and individual MiniDock MTB tests were reported.

The number of overall pools and specimens tested, the number of pools and specimens that tested positive, and the total number of tests performed were summarized. The observed testing efficiency was determined by dividing the number of assays used by the number of specimens with test results.

## RESULTS

From October to November 2025, 861 sputum swabs from 855 people were tested in 287 pools of 3, with 98 (11.4%) pooled swab test results positive, 752 (87.3%) negative and 11 (1.3%) inconclusive (**Figure 1**). Of the 1,097 participants included in the initial prospective study, 242 (22%) did not have stored sputum specimens available for inclusion in this secondary study. Among the 853 sputum swabs tested individually, 29 (3.4%) had invalid results (with no retesting).

Of the 855 participants with specimens included, 100 (12%) had culture-positive TB (**Table 1**).

**Table 1.** Participant demographic and clinical characteristics.

| Characteristic | Total |  |
| --- | --- | --- |
| Total | 855 |  |
| Age, years (median, IQR) | 49 | (36,64) |
| Sex |  |  |
| Female | 377 | (44) |
| Male | 478 | (56) |
| People living with HIV<sup>a</sup> | 153 | (18) |
| Cough > 2 weeks | 409 | (48) |
| TB symptoms, any of W4SS | 736 | (86) |
| Population |  |  |
| Health facility | 553 | (65) |
| Community | 302 | (35) |
| Sputum Xpert Ultra positive | 124 | (15) |
| Sputum TB culture positive<sup>b</sup> | 100 | (12) |
| Sputum smear-positive | 69 | (69) |
| Sputum smear-negative | 29 | (29) |

Abbreviations: W4SS: WHO-recommended four symptom screen for TB (cough, fever, night sweats, weight loss); IQR, interquartile range
<sup>a</sup>Also 75 people with status unknown and 629 with negative results
<sup>b</sup>2 with missing smear results

### Diagnostic performance of pooled testing in pools of three sputum swabs on the MiniDock MTB

Sensitivity of pooled testing in pools of three sputum swabs and individual sputum swab testing on the MiniDock MTB against the MRS was 88% (87/99, 95% CI, 80-93%) and 89% (88/99, 95% CI, 81-94%), respectively (**Table 2**). Specificity of pooled testing with three sputum swabs was 99% (98-99%) versus 95% (642/673, 94-97%) for individual sputum swab testing.

**Table 2.** Diagnostic accuracy of testing in pools of three sputum swabs and individual sputum swab testing on Pluslife MiniDock MTB, sputum on Xpert MTB/RIF Ultra, and sputum smear microscopy, as compared with the reference standard of TB culture.

|  | N<sup>a</sup> | Sensitivity, %<br>(95% CI, n/N) | Specificity<sup>c</sup><br>%<br>(95% CI, n/N) |
| --- | --- | --- | --- |
| Sputum swab - pooled MiniDock MTB (3:1) | 795 | 88% (80-93%)<br>87/99 | 99% (98-99%)<br>687/696 |
| Sputum swab - individual MiniDock MTB | 772 | 89% (81-94%)<br>88/99 | 95% (94-97%)<br>642/673 |
| Sputum - Xpert MTB/RIF Ultra<sup>d</sup> | 805 | 96% (90-98%)<br>96/100 | 97% (95-98%)<br>684/705 |
| Sputum smear microscopy<sup>e</sup> | 98 | 70% (61-79%)<br>69/98 | - |

Abbreviation: TB, tuberculosis.
<sup>a</sup>Number of specimens with valid results on culture and index or comparator test.
<sup>b</sup>1 individual and pooled swab result with no result (not tested), 2 missing smear results
<sup>c</sup>9 inconclusive pooled swab test results (6 invalid and 3 with no results); 32 inconclusive individual swab test results (25 invalid and 7 with no results); on these stored sputum specimens (with no re-testing)
<sup>d</sup>Ultra testing was performed on a different sputum specimen from the same participant (Figure S1); final Ultra result (after re-testing for invalid initial results)
<sup>e</sup>Only for participants with culture positive TB; among 100 culture positive participants, 2 with missing smear results

As compared to sputum testing on Ultra (on a different sputum specimen from the participant), the positive percent agreement was 75% (92/122, 67-82%) for testing sputum swabs in pools of three and 79% (95/121, 95% CI, 70-85%) for individual sputum swab testing on the MiniDock MTB; negative percent agreement was 99% (95% CI, 98-100%) and 96% (95% CI, 94-97%), respectively (**Table S1**).

As compared to individual testing on MiniDock MTB, the positive percent agreement of testing sputum swabs in pools of three was 80% (98/122, 95% CI, 72-86%, **Table S2**). Of the 24 specimens that were positive for TB on initial individual MiniDock MTB testing, 1 had a positive result on culture, and 3 had positive results on Xpert MTB/RIF Ultra; none of these had a positive result on the repeat MiniDock MTB test on the same specimen or on the other sputum specimen (**Table S3**).

### Pooled testing results and efficiency

A total of 287 pools of 3 sputum swabs were tested on the MiniDock MTB assay, including 91 pools (32%) with positive results and 4 pools (1.4%) with an invalid result. Of the 850 specimens with valid results, 98 specimens (11.5%) had positive results for TB (**Table 3**). Among the 91 positive pools, 81 (89%) were complete, with valid results for all individual specimens, and among these 55 (68%) had a single individual positive, 14 (17%) had 2 individual positives and 2 (2.5%) had 3 individual positives; 10 (12%) had no individual positive results. Among the 10 incomplete positive pools there were 8 pools with a single individual positive specimen; among the 4 pools with invalid results, there was a single individual positive specimen.

**Table 3.** Testing statistics and estimated efficiency of specimen pooling with the MiniDock MTB in pools of three sputum swabs with positivity of 12%.

| **Summary statistics- Pooled testing** |  |
| --- | --- |
| Number of pools with results | 287 |
| Number of pools with results of MTB detected | 91 |
| Number of pools with results of invalid | 4 |
| Number of specimens with valid test results produced<sup>a</sup> | 850 |
| Number of specimens with positive pooled testing results | 98 |
| Positivity rate | 12% |
| Number of MiniDock MTB tests needed<sup>b,c</sup> | 578 |
| **Efficiency** |  |
| Number of tests run per person with result<br>(# of tests used / # of specimens with results) | 0.68 |
| Estimated proportion of tests saved with pooled testing<br>(as compared to individual testing)<sup>c</sup> | 32% |

<sup>a</sup>11 specimens had inconclusive pooled testing results (Figure 1)
<sup>b</sup>Includes one test for each pool and one test for individual testing of each specimen in any pool of three with result of MTB detected or invalid
<sup>c</sup>Includes re-testing individual tests with invalid results, assuming an invalid rate of 1%, as obtained previously with fresh sputum specimens on the MiniDock MTB [4]

Valid results were produced for 850 specimens using 578 MiniDock tests, or 0.68 tests per specimen with result. The estimated proportion of tests saved as compared to individual testing was 32%, assuming 1% invalid results that needed re-testing, as obtained with MiniDock MTB testing on fresh sputum previously [4].

## DISCUSSION

Our results are the first to document the performance of pooled testing on the MiniDock MTB assay. Pooled testing using three swabbed sputum specimens detected 87 of the 88 people with culture-positive TB that individual testing on MiniDock MTB detected and would have saved 32% of the test costs in our population, where 12% of people tested had culture positive TB. These results suggest that pooled testing on MiniDock MTB is feasible and could be implemented to lower reagent costs for TB programs. While early studies of pooled testing using Xpert MTB/RIF and Ultra showed good performance in a range of settings against individual Ultra testing [23–26], they did not use culture, the preferred reference standard for the detection of TB. In this diagnostic accuracy study of pooled testing with the MiniDock MTB assay, culture is the reference standard with Ultra as a comparator test, providing robust evidence for this diagnostic accuracy evaluation.

In this study, we pooled three sputum swabs directly into the Pluslife Releasing Agent tube, at the elution step of the assay, and specimen dilution is not expected. However, a limitation is the amount of specimen (in this case sputum), that can be added to the tube. We found that with the addition of four sputum swabs (roughly 0.8mL sputum), more invalid results were obtained. Assuming the same positivity (11.5%) and an invalid rate of 6.3% (as measured by us initially on 64 pools of 4 samples), then pooling with pools of 4 would have resulted in similar test savings (approximately 33%) as reported here for pools of 3 with 1% invalid results.

Similar to the large multicentre study on pooled testing for Ultra [7], we found improved specificity for pooled testing against individual testing when using culture as a reference standard. Some individual Ultra and MiniDock MTB tests detected TB in specimens from people who were culture negative, while pooled testing on these specimens led to negative results. For pooled testing on Ultra, specimen dilution presumably contributes to reducing the amount of bacterial DNA present in the pool and leading to a negative pool result while the individual test is positive, for specimens with lower concentrations of bacteria. For pooled MiniDock MTB swab testing as described here, specimen dilution is not expected, but it is notable that there were 10 pools (3.4% of 287 total pools) that were positive, with no individual swab positive on re-testing, and conversely there were 24 individual swabs (4.0%) with positive results among 588 individual tests conducted on specimens tested in negative pools. It is unclear if these discordant results are linked to people with active TB; among the 24 positive individual swabs from negative pools, only 3 had either a positive culture or Ultra result. Discordant results on pooled testing, with positive pool results followed by negative results on all individual tests from the pool, have been reported previously with pooled testing, including 14.5% in pooled testing for cCMV [19], 2.5-5.3% in pooled testing for SARS-CoV-2 [27] and 3.6-7.8% for TB testing with Ultra [15,16]. These discordant results are likely in part attributable to assay variability, as has been reported with nucleic acid amplification tests generally, and this can contribute to both individual positive results with negative pool results and vice versa. Assay variability has been described in detail for PCR testing, due to variations in chemical efficiencies (of the enzymes, primer and/or template) and factors including detection, temperature and reagent volume [28,29]. While similar analyses have not yet been reported for the RHAM (RNase Hybridization-Assisted amplification) isothermal amplification method used in the MiniDock MTB [30], it is likely that similar factors will contribute to variability in this assay. Just as for pooled testing on Ultra testing, the clinical impact of discordant pools can be mitigated by following up all individuals with negative results from positive pools for repeat testing and ensuring each person either gets better or is re-evaluated for TB.

In addition to assay cost savings, the higher efficiency of pooled testing as compared to individual testing when positivity rates are relatively low has the major advantage of saving time for testing, which can help to reduce time to patient results. This could be of significant value in settings where large numbers of people are being tested for TB and fast results could facilitate immediate linkage to care, such as in community or prison settings.

This study had several limitations. Testing was conducted on stored sputum specimens rather than on fresh sputum; the proportion of results that were invalid on initial testing was higher for stored sputum than has been reported for testing on fresh sputum swabs (3.4% vs 1.0%)[4], perhaps due to degradation of the internal control during storage. Testing was performed on a different specimen than the Ultra comparator, so direct analysis against the semi-quantitative Ultra result was not conducted. We evaluated only pools of three sputum swabs, and future investigations can explore other pools sizes and specimens.

In summary, use of pooled testing of three sputum swabs on the Pluslife MiniDock MTB assay had similar sensitivity and saved 32% of tests as compared to individual sputum swab testing, suggesting that this approach could help to further expand access to molecular testing for TB, while also saving test reagents, personnel time, equipment usage, and reducing time to diagnosis. Further studies are needed on the performance of this approach in prospective designs with a range of pool sizes and in different patient populations.

## Data Availability

All analyzed data are included in the supplementary information files.

## Notes

## Acknowledgments

We thank all participants for their contribution to this work.

We gratefully acknowledge all the contributions of the members of the RAPID TB team: Maurice Ganava Toussaint (National TB Program – Far North Region, Maroua, Cameroon), Ousmanou Bello (National TB Program – North Region, Garoua, Cameroon), Meoto Paul (National TB Program – Southwest Region, Buea, Cameroon), Fitame Adeline (National TB Program – West Region, Bafoussam, Cameroon), Pride Teyim (Tuberculosis Reference Laboratory Douala, Douala, Cameroon), Valerie Flore Donkeng-Donfack (Centre Pasteur du Cameroun); Nina Lubeka (Bonaberi Baptist Hospital, Bonaberi, Cameroon); Armand Koudjou (Bafoussam Baptist Hospital, Bafoussam, Cameroon); Guy Zero Molesa (Mboppi Baptist Hospital, Mboppi Cameroon); Joel Wepngong Tabah, Ngowo Eyambe Lydia (Mutengene Baptist Hospital, Mutengene, Cameroon); Christabelle Ewane (Bamenda Regional Hospital): Nguifu Kelly Ngwafung, Ambo Noeline Niba, Kolla Magang Nelly Celestine, Bechesi Carine, Boum Delphine Agnes Gaetan, Ngu Amanda Mambo, Yinkfu Marcel Ndamnsah, Munsi Valerie Nformi, Hella Adzoyo Reine, Elimbi Neline Yameni, Ahmadou Djenabou, Maiyanpa Moksala Josephine, Nchu-Nfor Evangeline, Iya A Nyam Aicha, Laban Rhoda Bongshe, Ngapgue Sobdong Dominique Josiane, Tsimene Tamessuing Sandrine, Gingir Beatrice Ndemnwi, Amaya Alhadji Wakar, Souk-ino Wappou Ferdinand, Guidzavai Koliye Paul, Naindouba Beninga Herve Steve, Eban Odi Luisa Nang, Tchindo Dargeo, Edzengte Pascal Steve Landry, Elisée Avaikdepainani, Njikam Daouda Momgbet, Mbah Romarick, Baiguerel Erika Myriam, Taya Fokou Jean Bosco, Rita Nsamenang, Gildas Nguimfack, Joceline Konso, Zourriyah Adamou Mana, Nankouo Arthur, Tiamuh Nzebele Magdalene(Center for Health Promotion and Research)

## Data sharing statement

All analyzed data are included in the supplementary information files.

