## Supplemental Materials for "Diagnostic accuracy and testing efficiency of pooled testing of sputum swabs to detect tuberculosis on the near point of care Pluslife MiniDock MTB assay"

**Supplemental File 1**

**Online supplement to Diagnostic accuracy and testing efficiency of pooled testing of sputum swabs to detect tuberculosis on the near point of care PlusLife MiniDock MTB assay**

Nadege Bonkar Chifu[1]*, Asonganyi Etiendem[1], Diana Kolieghu Tcheumeni[1], Angela Neh[1], Ngha Ndze Mbuh[1], Gilford Fonyuy[1], Denis Nsame[2], Norah Nyah Ndi[3], Irene Adeline Goupeyou Wandji[4], Mercy Fundoh[5], Cyrille Mbuli[1], Nestor Biatu[1], Comfort Vuchas[1], Tushar Garg[6], Jacob Creswell[6], Melissa Sander[1] and the RAPID TB team**

**Affiliated organization**

[1]Center for Health Promotion and Research, Bamenda, Cameroon
[2]Bamenda Regional Hospital, Bamenda, Cameroon
[3]Cameroon Baptist Convention Health Services, Bamenda, Cameroon
[4]National TB Program – Littoral Region, Douala, Cameroon
[5]National TB Program – Northwest Region, Bamenda, Cameroon
[6]Stop TB Partnership, Geneva, Switzerland

**Supplemental Methods**

**Study design and population**

This was a diagnostic accuracy study to assess pooled testing on the PlusLife MiniDock MTB assay using stored sputum specimens collected from outpatients and attendees at community-based screening events as part of a prospective study conducted from February to June 2025 in Cameroon (4). At five health facilities, participants included consecutive outpatients aged 15 years and above with symptoms (current cough, fever, night sweats and/or weight loss) and/or clinical risk factors (diabetes, current smoker) for TB. At community-based active case finding (ACF), participants included people aged 15 years and above who were screened for TB using chest X-ray with AI (qXR V4, Qure.ai, Mumbai, India) and had an AI score of ≥0.30. Individuals who were on TB treatment within the previous six months were excluded.

***Pool size determination***

Testing on the MiniDock MTB has previously been reported with the use of single sputum swabs (i.e. one sputum swab from one person). Based on the positivity of 11% (114/996) MiniDock MTB sputum swabs in the original prospective study,(4) the optimal pool size following Dorfman is four.(23) To assess the possibility of performing pooled testing with multiple sputum swabs on one assay, we added larger volumes of sputum than normally used to the Nucleic Acid Releasing Agent (NARA) tube. At volumes greater than approximately 1mL of sputum, the assay generated a high proportion of invalid results. In our experience, sputum swabs for testing on the MiniDock MTB had an average volume of approximately 0.2mL, with smaller volumes from swabs prepared with more liquid specimens and larger swab volumes from more viscous specimens. Based on this, we initially started with pools of four sputum swabs (with an anticipated approximate volume of 0.8mL sputum added to the NARA tube), but the rate of invalid results was greater than 6% (4/64 = 6.3%). We therefore switched to pools of three sputum swabs for this evaluation.

***Reference standard and comparator tests***

The reference standard for this study was TB culture, and comparator tests were smear microscopy and Xpert MTB/RIF Ultra (Ultra), as recommended for evaluations of sputum-based tests to detect TB(24). In the original prospective cohort(4), participants provided two sputum specimens; smear microscopy was performed on both sputum specimens, MiniDock MTB and Ultra testing were performed on the first specimen, and the second specimen was sent to the reference laboratory for TB culture (Figure S1). TB culture and Ultra testing have been described previously (ref). The comparator Ultra assay produces results of invalid, error, MTB not detected, or MTB detected (with grades of high, medium, low, very low, or trace). Technicians performing testing were blinded to other TB test results and clinical information.

***Review of discordant results***

After analysis of the results, for specimens that tested positive on the MiniDock MTB and had negative results by the reference standard of culture, new sputum swabs were prepared from the remaining aliquot and tested a second time on the MiniDock MTB. In addition, these results were compared to the PlusLIfe and Ultra results from the first sputum specimen that was tested in the original prospective study.

**Supplemental Discussion**

While the current WHO guidelines recommend using pools of up to four samples for Ultra testing, there have been studies that have conducted pooled testing using 8 or more samples (14) and Cameroon has reported larger pool size testing in programmatic use (16). Future studies could explore pooled testing of alternative sample preparations to assess the feasibility of testing larger pool sizes with sputum on this assay. Alternative or modified pool preparation methods might be possible, for example, by: adding less sputum from each specimen (with smaller swabs or reduction of sputum from the swab after preparation), by pooling specimens after lysis rather than at the elution step, and/or by eluting a small number of sputum swabs (i.e. 2, 3, or 4) into each NARA tube and then combining the lysate from multiple NARA tubes prior to testing (to reduce dilution). While we tested sputum swabs, it is expected that pools of tongue swabs could be performed with many more swabs, since the amount of biological material collected per swab is much less than the approximately 0.2mL obtained with a sputum swab. Pooled testing on the MiniDock MTB requires purchase of additional swabs (beyond the one swab per tested that is provided in the test kit); for this study of sputum swabs, we purchased locally available swabs for 80CFA ($0.14) each.

Pooled testing has been used in public health programs for decades. In the TB community this has been driven by a need to effectively reduce the cost per person tested as national budgets are stretched thin, and because the need for molecular testing is great. Most countries are facing cuts to their TB program budgets, as Global Fund, the world's largest multilateral TB donor, has announced cuts for its next grant cycle, driven by drastically reduced support to global health from many donor countries. Erratic supply chain issues also contributed to the need for alternatives to individual testing, especially during the COVID-19 epidemic, but even without the pandemic, many countries have faced test shortages for Ultra. The near point of care MiniDock MTB assay has potential to be used in many more health facilities as it can be decentralized, facilitating greater access to molecular TB testing. However, decentralization also brings additional supply chain complexities, and new near point of care test suppliers may face issues with production capacity, local distribution networks, and regulatory challenges that are common for new products. Having the ability to employ pooled testing, if reagent shortages occur, could help TB programs provide continuous diagnostic services in spite of these challenges.

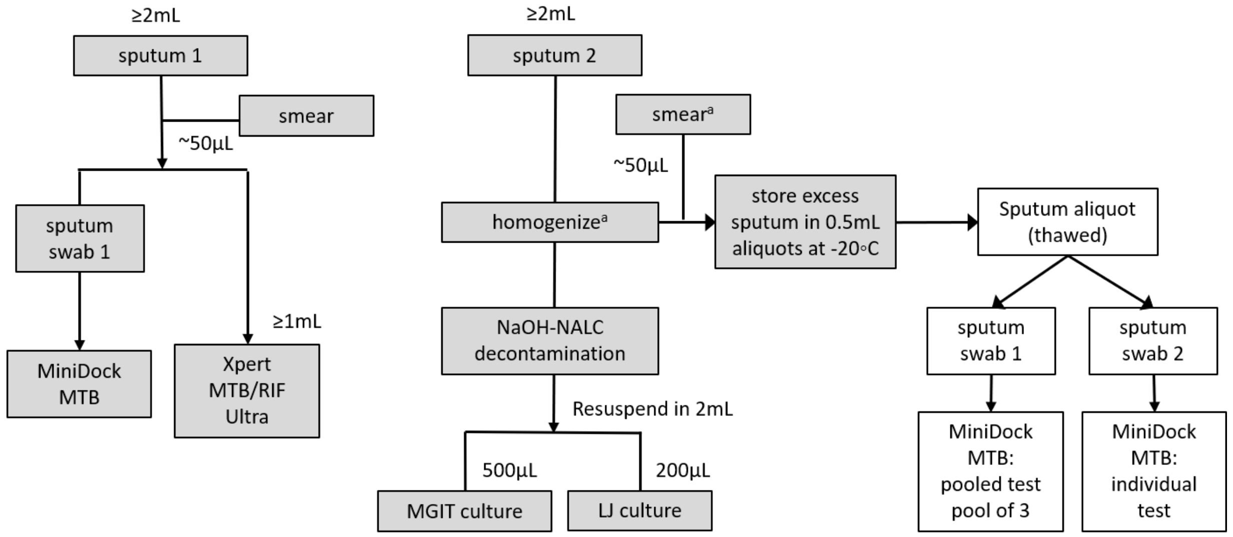

**Figure S1. Specimen flow.**

Steps performed in the original prospective study are shown in grey; steps performed in this secondary study on stored sputum are shown in white.

[a]For sputum specimens with volumes of 2mL (not excess), samples were not homogenized and smears were prepared directly.

**Table S1. Diagnostic agreement of sputum swabs in pools of three and individual sputum swabs on MiniDock MTB with sputum on Xpert MTB/RIF Ultra**

| | | Positive percent agreement | | | Negative percent agreement | | |
|---|---|---|---|---|---|---|---|
| | N[a] | % | (95% CI) | n/N | % | (95% CI) | n/N |
| Sputum swabs - pooled MiniDock MTB (3:1) [b] | 850 | 75% | (67-82%) | 92/122 | 99% | (98-100%) | 722/728 |
| Sputum swab - individual MiniDock MTB [b] | 824 | 79% | (70-85%) | 95/121 | 96% | (94-97%) | 676/703 |

Abbreviation: TB, tuberculosis.
[a]Number of specimens with valid results on culture and index or comparator test.
[b] 7 inconclusive pooled swab test results; 29 inconclusive individual swab test results; on these stored sputum specimens (with no re-testing)
Ultra testing was performed on a different sputum specimen from the same participant (Figure S1); final Ultra result (after re-testing for invalid initial results)

**Table S2. Positive percent agreement of sputum swabs in pools of three with individual sputum swabs on PlusLife MiniDock MTB**

| | Positive percent agreement | | |
|---|---|---|---|
| | % | (95% CI) | n/N |
| Sputum swabs - pooled MiniDock MTB (3:1) | 80% | (72-86%) | 98/122 |

**Table S3. Complete laboratory results for specimens that tested negative for TB on testing of three sputum swabs on the MiniDock MTB assay and positive for TB on an initial individual MiniDock MTB test.**

| Specimen # | Age Group | Sex | Sputum specimen #2 | | | | Sputum specimen #1 | |
|---|---|---|---|---|---|---|---|---|
| | | | MiniDock MTB results | | | Culture Result | Xpert MTB/RIF Ultra result | MiniDock MTB individual result |
| | | | Pool Test Result (3:1) | Individual result #1 | Individual result #2 | | | |
| 1 | 55-64 | M | Negative | **Positive** | Negative | Negative | Negative | Negative |
| 2 | 45-54 | F | Negative | **Positive** | Negative | Negative | Negative | Negative |
| 3 | 25-34 | M | Negative | **Positive** | Negative | Negative | Negative | Negative |
| 4 | 35-44 | M | Negative | **Positive** | Negative | Negative | Negative | Negative |
| 5 | 35-44 | M | Negative | **Positive** | Negative | Negative | Negative | Negative |
| 6 | 15-24 | F | Negative | **Positive** | Negative | Negative | Negative | Negative |
| 7 | 55-64 | F | Negative | **Positive** | Negative | Negative | Negative | Negative |
| 8 | 35-44 | F | Negative | **Positive** | Negative | Negative | Negative | Negative |
| 9 | 55-64 | F | Negative | **Positive** | Not done | Negative | Negative | Negative |
| 10 | 65+ | F | Negative | **Positive** | Not done | Negative | Negative | Negative |
| 11 | 65+ | F | Negative | **Positive** | Negative | Negative | Negative | Negative |
| 12 | 65+ | F | Negative | **Positive** | Negative | Negative | Negative | Negative |
| 13 | 15-24 | F | Negative | **Positive** | Negative | Contaminated | **Positive, Very low** | Negative |
| 14 | 65+ | M | Negative | **Positive** | Negative | Negative | Negative | Negative |
| 15 | 55-64 | M | Negative | **Positive** | Negative | Negative | Negative | Negative |
| 16 | 15-24 | F | Negative | **Positive** | Negative | Negative | Negative | Negative |
| 17 | 45-54 | M | Negative | **Positive** | Negative | Negative | Negative | Negative |
| 18 | 25-34 | M | Negative | **Positive** | Negative | Negative | Negative | Negative |
| 19 | 25-34 | M | Negative | **Positive** | Negative | **Positive** | **Positive, Trace** | Negative |
| 20 | 55-64 | F | Negative | **Positive** | Negative | Negative | Negative | Negative |
| 21 | 45-54 | M | Negative | **Positive** | Negative | Negative | **Positive, Trace** | Negative |
| 22 | 55-64 | F | Negative | **Positive** | Negative | Negative | Negative | Negative |
| 23 | 45-54 | M | Negative | **Positive** | Negative | Negative | Negative | Negative |
| 24 | 45-54 | M | Negative | **Positive** | Negative | Negative | Negative | Negative |
