## Supplemental Data for "Diagnostic accuracy and testing efficiency of pooled testing of sputum swabs to detect tuberculosis on the near point of care Pluslife MiniDock MTB assay"

| | pool_num | study_num | pool_result | indiv_resul | spec_pool_ | culture_res | Ultra_result_spec1 |
|---|---|---|---|---|---|---|---|
| 1 | 1 | 4 | Negative | Negative | Negative | 2-Negative | 2-Negative |
| 2 | 1 | 2 | Negative | Negative | Negative | 2-Negative | 2-Negative |
| 3 | 1 | 1 | Negative | Negative | Negative | 2-Negative | 2-Negative |
| 4 | 2 | 6 | Negative | Positive | Negative | 2-Negative | 2-Negative |
| 5 | 2 | 7 | Negative | Negative | Negative | 2-Negative | 2-Negative |
| 6 | 2 | 5 | Negative | Negative | Negative | 2-Negative | 2-Negative |
| 7 | 3 | 8 | Negative | Negative | Negative | 2-Negative | 2-Negative |
| 8 | 3 | 9 | Negative | Negative | Negative | 2-Negative | 2-Negative |
| 9 | 3 | 334 | Negative | Negative | Negative | 2-Negative | 2-Negative |
| 10 | 4 | 338 | Negative | Negative | Negative | 2-Negative | 2-Negative |
| 11 | 4 | 335 | Negative | Negative | Negative | 2-Negative | 2-Negative |
| 12 | 4 | 336 | Negative | Negative | Negative | 2-Negative | 2-Negative |
| 13 | 5 | 11 | Negative | Negative | Negative | 2-Negative | 2-Negative |
| 14 | 5 | 10 | Negative | Negative | Negative | 2-Negative | 2-Negative |
| 15 | 5 | 12 | Negative | Negative | Negative | 2-Negative | 2-Negative |
| 16 | 6 | 341 | Positive | Positive | Positive | 1-Positive | 1-Positive |
| 17 | 6 | 156 | Positive | Negative | Negative | 2-Negative | 2-Negative |
| 18 | 6 | 155 | Positive | Positive | Positive | 1-Positive | 1-Positive |
| 19 | 7 | 343 | Positive | Positive | Positive | 1-Positive | 1-Positive |
| 20 | 7 | 13 | Positive | Positive | Positive | 1-Positive | 1-Positive |
| 21 | 7 | 344 | Positive | Negative | Negative | 2-Negative | 2-Negative |
| 22 | 8 | 347 | Negative | Negative | Negative | 2-Negative | 2-Negative |
| 23 | 8 | 340 | Negative | Negative | Negative | 2-Negative | 2-Negative |
| 24 | 8 | 158 | Negative | Positive | Negative | 2-Negative | 2-Negative |
| 25 | 9 | 345 | Negative | Negative | Negative | 1-Positive | 2-Negative |
| 26 | 9 | 339 | Negative | Negative | Negative | 2-Negative | 2-Negative |
| 27 | 9 | 14 | Negative | Negative | Negative | 2-Negative | 2-Negative |
| 28 | 10 | 15 | Negative | Negative | Negative | 2-Negative | 2-Negative |
| 29 | 10 | 157 | Negative | Negative | Negative | 2-Negative | 2-Negative |
| 30 | 10 | 346 | Negative | Negative | Negative | 2-Negative | 2-Negative |
| 31 | 11 | 159 | Negative | Negative | Negative | 2-Negative | 2-Negative |
| 32 | 11 | 16 | Negative | Negative | Negative | 2-Negative | 2-Negative |
| 33 | 11 | 18 | Negative | Negative | Negative | 2-Negative | 2-Negative |
| 34 | 12 | 161 | Negative | Negative | Negative | 2-Negative | 2-Negative |
| 35 | 12 | 162 | Negative | Negative | Negative | 2-Negative | 2-Negative |
| 36 | 12 | 348 | Negative | Negative | Negative | 2-Negative | 2-Negative |
| 37 | 13 | 349 | Negative | Negative | Negative | 2-Negative | 2-Negative |
| 38 | 13 | 351 | Negative | Negative | Negative | 2-Negative | 2-Negative |
| 39 | 13 | 163 | Negative | Negative | Negative | 2-Negative | 2-Negative |
| 40 | 14 | 17 | Negative | Negative | Negative | 2-Negative | 2-Negative |
| 41 | 14 | 164 | Negative | Negative | Negative | 2-Negative | 2-Negative |
| 42 | 14 | 19 | Negative | Negative | Negative | 2-Negative | 2-Negative |
| 43 | 15 | 178 | Positive | Positive | Positive | 1-Positive | 1-Positive |
| 44 | 15 | 176 | Positive | Negative | Negative | 2-Negative | 2-Negative |
| 45 | 15 | 177 | Positive | Negative | Negative | 2-Negative | 2-Negative |
| 46 | 16 | 33 | Positive | Positive | Positive | 1-Positive | 1-Positive |
| 47 | 16 | 21 | Positive | Negative | Negative | 2-Negative | 2-Negative |

| | | | | | | | |
|---|---|---|---|---|---|---|---|
| 48 | 16 | 362 | Positive | Negative | Negative | 2-Negative | 2-Negative |
| 49 | 17 | 352 | Positive | Negative | Negative | 2-Negative | 2-Negative |
| 50 | 17 | 350 | Positive | Positive | Positive | 1-Positive | 1-Positive |
| 51 | 17 | 165 | Positive | NoResult | NoResult | 2-Negative | 2-Negative |
| 52 | 18 | 167 | Negative | Negative | Negative | 2-Negative | 2-Negative |
| 53 | 18 | 354 | Negative | Positive | Negative | 2-Negative | 2-Negative |
| 54 | 18 | 353 | Negative | Negative | Negative | 2-Negative | 2-Negative |
| 55 | 19 | 170 | Negative | Negative | Negative | 2-Negative | 2-Negative |
| 56 | 19 | 168 | Negative | Negative | Negative | 2-Negative | 2-Negative |
| 57 | 19 | 36 | Negative | Negative | Negative | 2-Negative | 2-Negative |
| 58 | 20 | 355 | Positive | Positive | Positive | 1-Positive | 1-Positive |
| 59 | 20 | 38 | Positive | Negative | Negative | 2-Negative | 2-Negative |
| 60 | 20 | 20 | Positive | Negative | Negative | 2-Negative | 2-Negative |
| 61 | 21 | 24 | Negative | Negative | Negative | 2-Negative | 2-Negative |
| 62 | 21 | 26 | Negative | Negative | Negative | 2-Negative | 2-Negative |
| 63 | 21 | 27 | Negative | Negative | Negative | 2-Negative | 2-Negative |
| 64 | 22 | 25 | Negative | Negative | Negative | 2-Negative | 2-Negative |
| 65 | 22 | 23 | Negative | Negative | Negative | 2-Negative | 2-Negative |
| 66 | 22 | 356 | Negative | Negative | Negative | 2-Negative | 2-Negative |
| 67 | 23 | 358 | Negative | Negative | Negative | 2-Negative | 2-Negative |
| 68 | 23 | 46 | Negative | Negative | Negative | 2-Negative | 2-Negative |
| 69 | 23 | 28 | Negative | Negative | Negative | 2-Negative | 2-Negative |
| 70 | 24 | 172 | Positive | Negative | Negative | 2-Negative | 2-Negative |
| 71 | 24 | 29 | Positive | Positive | Positive | 1-Positive | 1-Positive |
| 72 | 24 | 173 | Positive | Positive | Positive | 1-Positive | 1-Positive |
| 73 | 25 | 360 | Positive | NoResult | NoResult | 2-Negative | 2-Negative |
| 74 | 25 | 174 | Positive | Negative | Negative | 2-Negative | 2-Negative |
| 75 | 25 | 175 | Positive | Positive | Positive | 2-Negative | 1-Positive |
| 76 | 26 | 361 | Negative | Negative | Negative | 2-Negative | 2-Negative |
| 77 | 26 | 30 | Negative | Negative | Negative | 2-Negative | 2-Negative |
| 78 | 26 | 31 | Negative | Negative | Negative | 2-Negative | 2-Negative |
| 79 | 27 | 179 | Invalid | Negative | Negative | 2-Negative | 2-Negative |
| 80 | 27 | 181 | Invalid | Negative | Negative | 2-Negative | 2-Negative |
| 81 | 27 | 180 | Invalid | Negative | Negative | 2-Negative | 1-Positive |
| 82 | 28 | 182 | Positive | Negative | Negative | 2-Negative | 2-Negative |
| 83 | 28 | 420 | Positive | Positive | Positive | 1-Positive | 1-Positive |
| 84 | 28 | 22 | Positive | Negative | Negative | 2-Negative | 2-Negative |
| 85 | 29 | 421 | Positive | Negative | Negative | 2-Negative | 2-Negative |
| 86 | 29 | 423 | Positive | Positive | Positive | 1-Positive | 1-Positive |
| 87 | 29 | 34 | Positive | Negative | Negative | 2-Negative | 2-Negative |
| 88 | 30 | 39 | Negative | Negative | Negative | 2-Negative | 2-Negative |
| 89 | 30 | 37 | Negative | Negative | Negative | 2-Negative | 2-Negative |
| 90 | 30 | 183 | Negative | Negative | Negative | 2-Negative | 2-Negative |
| 91 | 31 | 169 | Negative | Negative | Negative | 2-Negative | 2-Negative |
| 92 | 31 | 368 | Negative | Negative | Negative | 2-Negative | 2-Negative |
| 93 | 31 | 367 | Negative | Negative | Negative | 2-Negative | 2-Negative |
| 94 | 32 | 422 | Positive | Negative | Negative | 2-Negative | 2-Negative |
| 95 | 32 | 431 | Positive | Positive | Positive | 1-Positive | 1-Positive |

| | | | | | | | |
|---|---|---|---|---|---|---|---|
| 96 | 32 | 170 | Positive | Negative | Negative | 2-Negative | 2-Negative |
| 97 | 33 | 425 | Positive | Negative | Negative | 2-Negative | 2-Negative |
| 98 | 33 | 424 | Positive | Positive | Positive | 1-Positive | 1-Positive |
| 99 | 33 | 184 | Positive | Negative | Negative | 2-Negative | 2-Negative |
| 100 | 34 | 185 | Positive | Invalid | Invalid | 2-Negative | 2-Negative |
| 101 | 34 | 363 | Positive | Negative | Negative | 2-Negative | 2-Negative |
| 102 | 34 | 364 | Positive | Positive | Positive | 1-Positive | 1-Positive |
| 103 | 35 | 186 | Positive | Positive | Positive | 1-Positive | 1-Positive |
| 104 | 35 | 366 | Positive | Negative | Negative | 2-Negative | 2-Negative |
| 105 | 35 | 32 | Positive | Negative | Negative | 2-Negative | 2-Negative |
| 106 | 36 | 193 | Negative | Negative | Negative | 2-Negative | 2-Negative |
| 107 | 36 | 194 | Negative | Negative | Negative | 2-Negative | 2-Negative |
| 108 | 36 | 426 | Negative | Negative | Negative | 2-Negative | 2-Negative |
| 109 | 37 | 196 | Negative | Positive | Negative | 2-Negative | 2-Negative |
| 110 | 37 | 195 | Negative | Negative | Negative | 2-Negative | 2-Negative |
| 111 | 37 | 370 | Negative | Negative | Negative | 2-Negative | 2-Negative |
| 112 | 38 | 372 | Negative | Positive | Negative | 2-Negative | 2-Negative |
| 113 | 38 | 373 | Negative | Negative | Negative | 2-Negative | 2-Negative |
| 114 | 38 | 371 | Negative | Negative | Negative | 2-Negative | 2-Negative |
| 115 | 39 | 374 | Positive | Negative | Negative | 2-Negative | 2-Negative |
| 116 | 39 | 375 | Positive | Negative | Negative | 2-Negative | 2-Negative |
| 117 | 39 | 435 | Positive | Positive | Positive | 1-Positive | 1-Positive |
| 118 | 40 | 436 | Positive | Positive | Positive | 1-Positive | 1-Positive |
| 119 | 40 | 377 | Positive | Positive | Positive | 1-Positive | 1-Positive |
| 120 | 40 | 376 | Positive | Negative | Negative | 2-Negative | 2-Negative |
| 121 | 41 | 197 | Positive | Negative | Negative | 2-Negative | 2-Negative |
| 122 | 41 | 434 | Positive | Positive | Positive | 1-Positive | 1-Positive |
| 123 | 41 | 56 | Positive | Negative | Negative | 1-Positive | 2-Negative |
| 124 | 42 | 199 | Negative | Positive | Negative | 2-Negative | 2-Negative |
| 125 | 42 | 200 | Negative | Negative | Negative | 2-Negative | 2-Negative |
| 126 | 42 | 198 | Negative | Negative | Negative | 2-Negative | 2-Negative |
| 127 | 43 | 61 | Negative | Negative | Negative | 2-Negative | 2-Negative |
| 128 | 43 | 58 | Negative | Negative | Negative | 2-Negative | 2-Negative |
| 129 | 43 | 57 | Negative | Negative | Negative | 2-Negative | 2-Negative |
| 130 | 44 | 41 | Negative | Negative | Negative | 2-Negative | 2-Negative |
| 131 | 44 | 437 | Negative | Negative | Negative | 2-Negative | 2-Negative |
| 132 | 44 | 59 | Negative | Negative | Negative | 2-Negative | 2-Negative |
| 133 | 45 | 438 | Negative | Negative | Negative | 2-Negative | 2-Negative |
| 134 | 45 | 62 | Negative | Negative | Negative | 2-Negative | 2-Negative |
| 135 | 45 | 63 | Negative | Negative | Negative | 2-Negative | 2-Negative |
| 136 | 46 | 65 | Positive | Negative | Negative | 2-Negative | 2-Negative |
| 137 | 46 | 68 | Positive | Negative | Negative | 2-Negative | 2-Negative |
| 138 | 46 | 63 | Positive | Negative | Negative | 2-Negative | 2-Negative |
| 139 | 47 | 202 | Positive | Negative | Negative | 2-Negative | 2-Negative |
| 140 | 47 | 201 | Positive | Positive | Positive | 1-Positive | 1-Positive |
| 141 | 47 | 69 | Positive | Negative | Negative | 2-Negative | 2-Negative |
| 142 | 48 | 439 | Negative | Negative | Negative | 2-Negative | 2-Negative |
| 143 | 48 | 378 | Negative | Positive | Negative | 2-Negative | 2-Negative |

| | | | | | | | |
|---|---|---|---|---|---|---|---|
| 144 | 48 | 440 | Negative | Negative | Negative | 2-Negative | 2-Negative |
| 145 | 49 | 442 | Negative | Negative | Negative | 2-Negative | 2-Negative |
| 146 | 49 | 441 | Negative | Positive | Negative | 2-Negative | 2-Negative |
| 147 | 49 | 443 | Negative | Negative | Negative | 2-Negative | 2-Negative |
| 148 | 50 | 428 | Negative | Negative | Negative | 2-Negative | 2-Negative |
| 149 | 50 | 427 | Negative | Negative | Negative | 2-Negative | 2-Negative |
| 150 | 50 | 429 | Negative | Negative | Negative | 2-Negative | 2-Negative |
| 151 | 51 | 44 | Positive | Positive | Positive | 1-Positive | 1-Positive |
| 152 | 51 | 43 | Positive | Negative | Negative | 2-Negative | 2-Negative |
| 153 | 51 | 430 | Positive | Negative | Negative | 2-Negative | 2-Negative |
| 154 | 52 | 188 | Positive | Negative | Negative | 2-Negative | 2-Negative |
| 155 | 52 | 45 | Positive | Positive | Positive | 1-Positive | 1-Positive |
| 156 | 52 | 187 | Positive | Negative | Negative | 2-Negative | 2-Negative |
| 157 | 53 | 369 | Positive | Positive | Positive | 1-Positive | 1-Positive |
| 158 | 53 | 49 | Positive | Negative | Negative | 2-Negative | 2-Negative |
| 159 | 53 | 48 | Positive | Negative | Negative | 2-Negative | 2-Negative |
| 160 | 54 | 192 | Negative | Invalid | Negative | 2-Negative | 2-Negative |
| 161 | 54 | 191 | Negative | Negative | Negative | 2-Negative | 2-Negative |
| 162 | 54 | 50 | Negative | Negative | Negative | 2-Negative | 2-Negative |
| 163 | 55 | 40 | Negative | Negative | Negative | 2-Negative | 2-Negative |
| 164 | 55 | 51 | Negative | Negative | Negative | 2-Negative | 2-Negative |
| 165 | 55 | 47 | Negative | Negative | Negative | 2-Negative | 2-Negative |
| 166 | 56 | 55 | Negative | Negative | Negative | 2-Negative | 2-Negative |
| 167 | 56 | 166 | Negative | Negative | Negative | 2-Negative | 2-Negative |
| 168 | 56 | 67 | Negative | Negative | Negative | 2-Negative | 2-Negative |
| 169 | 57 | 432 | Negative | Negative | Negative | 2-Negative | 2-Negative |
| 170 | 57 | 365 | Negative | Negative | Negative | 2-Negative | 2-Negative |
| 171 | 57 | 433 | Negative | Negative | Negative | 2-Negative | 2-Negative |
| 172 | 58 | 42 | Negative | Negative | Negative | 2-Negative | 2-Negative |
| 173 | 58 | 66 | Negative | Negative | Negative | 2-Negative | 2-Negative |
| 174 | 58 | 87 | Negative | Negative | Negative | 2-Negative | 2-Negative |
| 175 | 59 | 52 | Positive | Negative | Negative | 2-Negative | 2-Negative |
| 176 | 59 | 380 | Positive | Positive | Positive | 1-Positive | 1-Positive |
| 177 | 59 | 379 | Positive | Negative | Negative | 2-Negative | 2-Negative |
| 178 | 60 | 53 | Negative | Negative | Negative | 2-Negative | 2-Negative |
| 179 | 60 | 554 | Negative | Negative | Negative | 2-Negative | 2-Negative |
| 180 | 60 | 54 | Negative | Invalid | Negative | 2-Negative | 2-Negative |
| 181 | 61 | 557 | Negative | Negative | Negative | 2-Negative | 2-Negative |
| 182 | 61 | 555 | Negative | Negative | Negative | 2-Negative | 2-Negative |
| 183 | 61 | 556 | Negative | Negative | Negative | 2-Negative | 2-Negative |
| 184 | 62 | 558 | Positive | Negative | Negative | 1-Positive | 1-Positive |
| 185 | 62 | 559 | Positive | Negative | Negative | 2-Negative | 2-Negative |
| 186 | 62 | 560 | Positive | Negative | Negative | 2-Negative | 2-Negative |
| 187 | 63 | 562 | Positive | Invalid | Invalid | 2-Negative | 2-Negative |
| 188 | 63 | 561 | Positive | Positive | Positive | 1-Positive | 1-Positive |
| 189 | 63 | 566 | Positive | Negative | Negative | 2-Negative | 2-Negative |
| 190 | 64 | 565 | Negative | Negative | Negative | 2-Negative | 2-Negative |
| 191 | 64 | 563 | Negative | Negative | Negative | 2-Negative | 2-Negative |

| | | | | | | | |
|---|---|---|---|---|---|---|---|
| 192 | 64 | 564 | Negative | Negative | Negative | 2-Negative | 2-Negative |
| 193 | 65 | 569 | Negative | Negative | Negative | 2-Negative | 2-Negative |
| 194 | 65 | 567 | Negative | Negative | Negative | 2-Negative | 2-Negative |
| 195 | 65 | 568 | Negative | Invalid | Negative | 2-Negative | 2-Negative |
| 196 | 66 | 446 | Negative | Negative | Negative | 2-Negative | 2-Negative |
| 197 | 66 | 444 | Negative | Negative | Negative | 2-Negative | 2-Negative |
| 198 | 66 | 570 | Negative | Negative | Negative | 2-Negative | 2-Negative |
| 199 | 67 | 449 | Negative | Negative | Negative | 2-Negative | 2-Negative |
| 200 | 67 | 447 | Negative | Negative | Negative | 2-Negative | 2-Negative |
| 201 | 67 | 448 | Negative | Negative | Negative | 2-Negative | 2-Negative |
| 202 | 68 | 450 | Positive | Positive | Positive | 1-Positive | 1-Positive |
| 203 | 68 | 205 | Positive | Negative | Negative | 2-Negative | 2-Negative |
| 204 | 68 | 452 | Positive | Negative | Negative | 2-Negative | 2-Negative |
| 205 | 69 | 203 | Negative | Negative | Negative | 2-Negative | 2-Negative |
| 206 | 69 | 208 | Negative | NoResult | Negative | 2-Negative | 2-Negative |
| 207 | 69 | 206 | Negative | Negative | Negative | 2-Negative | 2-Negative |
| 208 | 70 | 382 | Negative | Negative | Negative | 2-Negative | 2-Negative |
| 209 | 70 | 209 | Negative | Negative | Negative | 2-Negative | 2-Negative |
| 210 | 70 | 210 | Negative | Negative | Negative | 2-Negative | 2-Negative |
| 211 | 71 | 207 | Negative | Negative | Negative | 2-Negative | 2-Negative |
| 212 | 71 | 71 | Negative | Negative | Negative | 2-Negative | 2-Negative |
| 213 | 71 | 212 | Negative | Negative | Negative | 2-Negative | 1-Positive |
| 214 | 72 | 79 | Negative | Negative | Negative | 2-Negative | 2-Negative |
| 215 | 72 | 211 | Negative | Negative | Negative | 2-Negative | 2-Negative |
| 216 | 72 | 213 | Negative | Negative | Negative | 2-Negative | 2-Negative |
| 217 | 73 | 80 | Negative | Negative | Negative | 2-Negative | 2-Negative |
| 218 | 73 | 81 | Negative | Negative | Negative | 2-Negative | 2-Negative |
| 219 | 73 | 82 | Negative | Negative | Negative | 2-Negative | 2-Negative |
| 220 | 74 | 577 | Negative | Negative | Negative | 2-Negative | 2-Negative |
| 221 | 74 | 83 | Negative | Negative | Negative | 2-Negative | 2-Negative |
| 222 | 74 | 204 | Negative | Negative | Negative | 2-Negative | 2-Negative |
| 223 | 75 | 576 | Negative | Negative | Negative | 2-Negative | 2-Negative |
| 224 | 75 | 578 | Negative | Invalid | Negative | 2-Negative | 2-Negative |
| 225 | 75 | 579 | Negative | Negative | Negative | 2-Negative | 1-Positive |
| 226 | 76 | 584 | Positive | Negative | Negative | 2-Negative | 2-Negative |
| 227 | 76 | 582 | Positive | Negative | Negative | 2-Negative | 1-Positive |
| 228 | 76 | 583 | Positive | Negative | Negative | 2-Negative | 2-Negative |
| 229 | 77 | 571 | Positive | Negative | Negative | 2-Negative | 2-Negative |
| 230 | 77 | 572 | Positive | Negative | Negative | 2-Negative | 2-Negative |
| 231 | 77 | 581 | Positive | Negative | Negative | 2-Negative | 2-Negative |
| 232 | 78 | 573 | Negative | Invalid | Negative | 2-Negative | 2-Negative |
| 233 | 78 | 575 | Negative | Invalid | Negative | 2-Negative | 2-Negative |
| 234 | 78 | 574 | Negative | Invalid | Negative | 2-Negative | 2-Negative |
| 235 | 79 | 229 | Positive | Negative | Negative | 2-Negative | 2-Negative |
| 236 | 79 | 228 | Positive | Negative | Negative | 2-Negative | 2-Negative |
| 237 | 79 | 389 | Positive | Positive | Positive | 1-Positive | 1-Positive |
| 238 | 80 | 453 | Positive | Negative | Negative | 2-Negative | 2-Negative |
| 239 | 80 | 230 | Positive | Positive | Positive | 1-Positive | 1-Positive |

| | | | | | | | |
|---|---|---|---|---|---|---|---|
| 240 | 80 | 383 | Positive | Invalid | Invalid | 2-Negative | 2-Negative |
| 241 | 81 | 454 | Positive | Negative | Negative | 2-Negative | 2-Negative |
| 242 | 81 | 385 | Positive | Positive | Positive | 1-Positive | 1-Positive |
| 243 | 81 | 90 | Positive | Negative | Negative | 2-Negative | 2-Negative |
| 244 | 82 | 70 | Negative | Negative | Negative | 2-Negative | 2-Negative |
| 245 | 82 | 458 | Negative | Negative | Negative | 2-Negative | 2-Negative |
| 246 | 82 | 216 | Negative | Positive | Negative | 2-Negative | 2-Negative |
| 247 | 83 | 451 | Negative | Negative | Negative | 2-Negative | 2-Negative |
| 248 | 83 | 445 | Negative | Negative | Negative | 2-Negative | 2-Negative |
| 249 | 83 | 243 | Negative | Negative | Negative | 2-Negative | 2-Negative |
| 250 | 84 | 456 | Negative | Negative | Negative | 2-Negative | 2-Negative |
| 251 | 84 | 78 | Negative | Positive | Negative | 2-Negative | 2-Negative |
| 252 | 84 | 84 | Negative | NoResult | Negative | 2-Negative | 2-Negative |
| 253 | 85 | 218 | Positive | Negative | Negative | 2-Negative | 2-Negative |
| 254 | 85 | 215 | Positive | Positive | Positive | 1-Positive | 1-Positive |
| 255 | 85 | 217 | Positive | Negative | Negative | 2-Negative | 2-Negative |
| 256 | 86 | 219 | Positive | Positive | Positive | 1-Positive | 1-Positive |
| 257 | 86 | 85 | Positive | Negative | Negative | 2-Negative | 2-Negative |
| 258 | 86 | 86 | Positive | Negative | Negative | 2-Negative | 2-Negative |
| 259 | 87 | 73 | Negative | Negative | Negative | 2-Negative | 2-Negative |
| 260 | 87 | 88 | Negative | Negative | Negative | 2-Negative | 2-Negative |
| 261 | 87 | 72 | Negative | Negative | Negative | 2-Negative | 2-Negative |
| 262 | 88 | 35 | Negative | Negative | Negative | 2-Negative | 2-Negative |
| 263 | 88 | 214 | Negative | Negative | Negative | 2-Negative | 2-Negative |
| 264 | 88 | 464 | Negative | Negative | Negative | 2-Negative | 2-Negative |
| 265 | 89 | 468 | Negative | Negative | Negative | 2-Negative | 2-Negative |
| 266 | 89 | 467 | Negative | Negative | Negative | 2-Negative | 2-Negative |
| 267 | 89 | 466 | Negative | Negative | Negative | 2-Negative | 2-Negative |
| 268 | 90 | 469 | Positive | Negative | Negative | 2-Negative | 2-Negative |
| 269 | 90 | 471 | Positive | Positive | Positive | 1-Positive | 1-Positive |
| 270 | 90 | 472 | Positive | Negative | Negative | 2-Negative | 2-Negative |
| 271 | 91 | 615 | Positive | Negative | Negative | 2-Negative | 2-Negative |
| 272 | 91 | 237 | Positive | Positive | Positive | 1-Positive | 1-Positive |
| 273 | 91 | 614 | Positive | Negative | Negative | 2-Negative | 2-Negative |
| 274 | 92 | 617 | Negative | Negative | Negative | 2-Negative | 2-Negative |
| 275 | 92 | 616 | Negative | Negative | Negative | 2-Negative | 2-Negative |
| 276 | 92 | 618 | Negative | Negative | Negative | 2-Negative | 2-Negative |
| 277 | 93 | 621 | Negative | Negative | Negative | 2-Negative | 2-Negative |
| 278 | 93 | 620 | Negative | Negative | Negative | 3-Contami | 2-Negative |
| 279 | 93 | 619 | Negative | Negative | Negative | 2-Negative | 2-Negative |
| 280 | 94 | 622 | Negative | Negative | Negative | 2-Negative | 2-Negative |
| 281 | 94 | 624 | Negative | Negative | Negative | 2-Negative | 2-Negative |
| 282 | 94 | 623 | Negative | Negative | Negative | 2-Negative | 2-Negative |
| 283 | 95 | 626 | Negative | Negative | Negative | 2-Negative | 2-Negative |
| 284 | 95 | 625 | Negative | Invalid | Negative | 2-Negative | 2-Negative |
| 285 | 95 | 627 | Negative | Negative | Negative | 2-Negative | 2-Negative |
| 286 | 96 | 606 | Negative | Invalid | Negative | 2-Negative | 2-Negative |
| 287 | 96 | 605 | Negative | Invalid | Negative | 2-Negative | 1-Positive |

| | | | | | | | |
|---|---|---|---|---|---|---|---|
| 288 | 96 | 628 | Negative | Invalid | Negative | 2-Negative | 2-Negative |
| 289 | 97 | 608 | Negative | Negative | Negative | 2-Negative | 2-Negative |
| 290 | 97 | 607 | Negative | Negative | Negative | 2-Negative | 1-Positive |
| 291 | 97 | 609 | Negative | Negative | Negative | 2-Negative | 2-Negative |
| 292 | 98 | 612 | Positive | Negative | Negative | 3-Contami | 2-Negative |
| 293 | 98 | 610 | Positive | Positive | Positive | 3-Contami | 1-Positive |
| 294 | 98 | 611 | Positive | Negative | Negative | 2-Negative | 2-Negative |
| 295 | 99 | 613 | Negative | Negative | Negative | 2-Negative | 2-Negative |
| 296 | 99 | 226 | Negative | NoResult | Negative | 2-Negative | 2-Negative |
| 297 | 99 | 75 | Negative | Negative | Negative | 2-Negative | 2-Negative |
| 298 | 100 | 93 | Negative | Negative | Negative | 2-Negative | 2-Negative |
| 299 | 100 | 224 | Negative | Positive | Negative | 2-Negative | 2-Negative |
| 300 | 100 | 223 | Negative | Negative | Negative | 2-Negative | 2-Negative |
| 301 | 101 | 587 | Positive | Negative | Negative | 2-Negative | 2-Negative |
| 302 | 101 | 460 | Positive | Positive | Positive | 1-Positive | 1-Positive |
| 303 | 101 | 586 | Positive | Negative | Negative | 1-Positive | 1-Positive |
| 304 | 102 | 590 | Negative | Negative | Negative | 2-Negative | 2-Negative |
| 305 | 102 | 589 | Negative | Negative | Negative | 2-Negative | 1-Positive |
| 306 | 102 | 588 | Negative | Negative | Negative | 2-Negative | 2-Negative |
| 307 | 103 | 593 | Positive | Negative | Negative | 3-Contami | 2-Negative |
| 308 | 103 | 591 | Positive | Positive | Positive | 2-Negative | 1-Positive |
| 309 | 103 | 592 | Positive | Negative | Negative | 2-Negative | 2-Negative |
| 310 | 104 | 594 | Negative | Negative | Negative | 2-Negative | 2-Negative |
| 311 | 104 | 459 | Negative | Negative | Negative | 2-Negative | 2-Negative |
| 312 | 104 | 89 | Negative | Negative | Negative | 2-Negative | 2-Negative |
| 313 | 105 | 597 | Negative | Invalid | Negative | 3-Contami | 2-Negative |
| 314 | 105 | 596 | Negative | Negative | Negative | 3-Contami | 2-Negative |
| 315 | 105 | 595 | Negative | Negative | Negative | 3-Contami | 1-Positive |
| 316 | 106 | 465 | Negative | Negative | Negative | 2-Negative | 2-Negative |
| 317 | 106 | 462 | Negative | Negative | Negative | 2-Negative | 2-Negative |
| 318 | 106 | 74 | Negative | Negative | Negative | 2-Negative | 2-Negative |
| 319 | 107 | 600 | Negative | Negative | Negative | 2-Negative | 1-Positive |
| 320 | 107 | 598 | Negative | Negative | Negative | 2-Negative | 2-Negative |
| 321 | 107 | 599 | Negative | Negative | Negative | 3-Contami | 2-Negative |
| 322 | 108 | 601 | Negative | Negative | Negative | 2-Negative | 2-Negative |
| 323 | 108 | 603 | Negative | Invalid | Negative | 3-Contami | 2-Negative |
| 324 | 108 | 602 | Negative | Negative | Negative | 2-Negative | 2-Negative |
| 325 | 109 | 94 | Positive | Negative | Negative | 2-Negative | 2-Negative |
| 326 | 109 | 76 | Positive | Negative | Negative | 2-Negative | 2-Negative |
| 327 | 109 | 604 | Positive | Positive | Positive | 1-Positive | 1-Positive |
| 328 | 110 | 97 | Invalid | Negative | Negative | 2-Negative | 2-Negative |
| 329 | 110 | 95 | Invalid | Negative | Negative | 2-Negative | 2-Negative |
| 330 | 110 | 96 | Invalid | Negative | Negative | 2-Negative | 2-Negative |
| 331 | 111 | 455 | Negative | Negative | Negative | 2-Negative | 2-Negative |
| 332 | 111 | 99 | Negative | Negative | Negative | 2-Negative | 2-Negative |
| 333 | 111 | 98 | Negative | Invalid | Negative | 2-Negative | 2-Negative |
| 334 | 112 | 457 | Negative | Negative | Negative | 2-Negative | 2-Negative |
| 335 | 112 | 386 | Negative | Negative | Negative | 2-Negative | 2-Negative |

| | | | | | | | |
|---|---|---|---|---|---|---|---|
| 336 | 112 | 222 | Negative | Negative | Negative | 2-Negative | 2-Negative |
| 337 | 113 | 388 | Negative | Negative | Negative | 2-Negative | 2-Negative |
| 338 | 113 | 387 | Negative | Negative | Negative | 2-Negative | 2-Negative |
| 339 | 113 | 92 | Negative | Negative | Negative | 2-Negative | 2-Negative |
| 340 | 114 | 233 | Positive | Negative | Negative | 2-Negative | 2-Negative |
| 341 | 114 | 232 | Positive | Positive | Positive | 1-Positive | 1-Positive |
| 342 | 114 | 231 | Positive | Negative | Negative | 2-Negative | 2-Negative |
| 343 | 115 | 235 | Negative | Negative | Negative | 2-Negative | 2-Negative |
| 344 | 115 | 236 | Negative | Negative | Negative | 2-Negative | 2-Negative |
| 345 | 115 | 234 | Negative | Negative | Negative | 2-Negative | 2-Negative |
| 346 | 116 | 110 | Negative | Negative | Negative | 2-Negative | 2-Negative |
| 347 | 116 | 109 | Negative | Negative | Negative | 2-Negative | 2-Negative |
| 348 | 116 | 240 | Negative | Negative | Negative | 2-Negative | 2-Negative |
| 349 | 117 | 227 | Negative | Negative | Negative | 2-Negative | 2-Negative |
| 350 | 117 | 77 | Negative | Negative | Negative | 2-Negative | 2-Negative |
| 351 | 117 | 104 | Negative | Negative | Negative | 2-Negative | 2-Negative |
| 352 | 118 | 101 | Negative | Negative | Negative | 2-Negative | 2-Negative |
| 353 | 118 | 103 | Negative | Negative | Negative | 2-Negative | 2-Negative |
| 354 | 118 | 102 | Negative | Negative | Negative | 2-Negative | 2-Negative |
| 355 | 119 | 239 | Negative | Negative | Negative | 2-Negative | 2-Negative |
| 356 | 119 | 100 | Negative | Negative | Negative | 2-Negative | 2-Negative |
| 357 | 119 | 391 | Negative | Negative | Negative | 2-Negative | 2-Negative |
| 358 | 120 | 473 | Negative | Negative | Negative | 2-Negative | 2-Negative |
| 359 | 120 | 478 | Negative | Negative | Negative | 3-Contami | 2-Negative |
| 360 | 120 | 474 | Negative | Negative | Negative | 2-Negative | 2-Negative |
| 361 | 121 | 394 | Positive | Positive | Positive | 1-Positive | 1-Positive |
| 362 | 121 | 475 | Positive | Negative | Negative | 2-Negative | 2-Negative |
| 363 | 121 | 476 | Positive | Negative | Negative | 2-Negative | 2-Negative |
| 364 | 122 | 512 | Positive | Negative | Negative | 2-Negative | 2-Negative |
| 365 | 122 | 241 | Positive | Positive | Positive | 1-Positive | 1-Positive |
| 366 | 122 | 393 | Positive | Negative | Negative | 2-Negative | 2-Negative |
| 367 | 123 | 470 | Positive | Positive | Positive | 1-Positive | 1-Positive |
| 368 | 123 | 514 | Positive | Negative | Negative | 2-Negative | 2-Negative |
| 369 | 123 | 511 | Positive | Negative | Negative | 2-Negative | 2-Negative |
| 370 | 124 | 513 | Negative | Negative | Negative | 2-Negative | 2-Negative |
| 371 | 124 | 107 | Negative | Negative | Negative | 2-Negative | 2-Negative |
| 372 | 124 | 106 | Negative | Negative | Negative | 2-Negative | 2-Negative |
| 373 | 125 | 238 | Positive | Negative | Negative | 2-Negative | 2-Negative |
| 374 | 125 | 105 | Positive | Negative | Negative | 2-Negative | 2-Negative |
| 375 | 125 | 392 | Positive | Positive | Positive | 1-Positive | 1-Positive |
| 376 | 126 | 480 | Negative | Negative | Negative | 2-Negative | 2-Negative |
| 377 | 126 | 479 | Negative | Negative | Negative | 2-Negative | 2-Negative |
| 378 | 126 | 477 | Negative | Negative | Negative | 2-Negative | 2-Negative |
| 379 | 127 | 521 | Negative | Negative | Negative | 2-Negative | 2-Negative |
| 380 | 127 | 515 | Negative | Negative | Negative | 2-Negative | 2-Negative |
| 381 | 127 | 516 | Negative | Negative | Negative | 2-Negative | 2-Negative |
| 382 | 128 | 519 | Negative | Negative | Negative | 2-Negative | 2-Negative |
| 383 | 128 | 518 | Negative | Negative | Negative | 2-Negative | 2-Negative |

| | | | | | | | |
|---|---|---|---|---|---|---|---|
| 384 | 128 | 520 | Negative | Negative | Negative | 2-Negative | 2-Negative |
| 385 | 129 | 517 | Positive | Negative | Negative | 2-Negative | 2-Negative |
| 386 | 129 | 671 | Positive | Positive | Positive | 1-Positive | 1-Positive |
| 387 | 129 | 481 | Positive | Negative | Negative | 2-Negative | 2-Negative |
| 388 | 130 | 672 | Negative | Negative | Negative | 1-Positive | 1-Positive |
| 389 | 130 | 673 | Negative | Invalid | Negative | 2-Negative | 2-Negative |
| 390 | 130 | 674 | Negative | Negative | Negative | 2-Negative | 1-Positive |
| 391 | 131 | 677 | Negative | Negative | Negative | 2-Negative | 2-Negative |
| 392 | 131 | 676 | Negative | Negative | Negative | 2-Negative | 2-Negative |
| 393 | 131 | 675 | Negative | Negative | Negative | 2-Negative | 2-Negative |
| 394 | 132 | 678 | Negative | Negative | Negative | 2-Negative | 2-Negative |
| 395 | 132 | 679 | Negative | Negative | Negative | 2-Negative | 2-Negative |
| 396 | 132 | 680 | Negative | Negative | Negative | 3-Contami | 2-Negative |
| 397 | 133 | 682 | Positive | Negative | Negative | 2-Negative | 2-Negative |
| 398 | 133 | 681 | Positive | Negative | Negative | 3-Contami | 2-Negative |
| 399 | 133 | 683 | Positive | Negative | Negative | 2-Negative | 2-Negative |
| 400 | 134 | 685 | Negative | Negative | Negative | 2-Negative | 2-Negative |
| 401 | 134 | 686 | Negative | Negative | Negative | 2-Negative | 2-Negative |
| 402 | 134 | 684 | Negative | Negative | Negative | 2-Negative | 2-Negative |
| 403 | 135 | 688 | Positive | Negative | Negative | 2-Negative | 2-Negative |
| 404 | 135 | 689 | Positive | Negative | Negative | 1-Positive | 1-Positive |
| 405 | 135 | 687 | Positive | Negative | Negative | 3-Contami | 2-Negative |
| 406 | 136 | 690 | Positive | Negative | Negative | 3-Contami | 2-Negative |
| 407 | 136 | 692 | Positive | Negative | Negative | 1-Positive | 1-Positive |
| 408 | 136 | 691 | Positive | Positive | Positive | 1-Positive | 1-Positive |
| 409 | 137 | 693 | Negative | Negative | Negative | 3-Contami | 2-Negative |
| 410 | 137 | 695 | Negative | Negative | Negative | 2-Negative | 2-Negative |
| 411 | 137 | 694 | Negative | Negative | Negative | 2-Negative | 2-Negative |
| 412 | 138 | 696 | Negative | Negative | Negative | 2-Negative | 2-Negative |
| 413 | 138 | 697 | Negative | Negative | Negative | 2-Negative | 2-Negative |
| 414 | 138 | 698 | Negative | Negative | Negative | 2-Negative | 2-Negative |
| 415 | 139 | 700 | Negative | Negative | Negative | 3-Contami | 2-Negative |
| 416 | 139 | 484 | Negative | Positive | Negative | 2-Negative | 2-Negative |
| 417 | 139 | 699 | Negative | Negative | Negative | 2-Negative | 1-Positive |
| 418 | 140 | 645 | Negative | Negative | Negative | 3-Contami | 2-Negative |
| 419 | 140 | 647 | Negative | Negative | Negative | 3-Contami | 2-Negative |
| 420 | 140 | 646 | Negative | Negative | Negative | 2-Negative | 2-Negative |
| 421 | 141 | 648 | Positive | Negative | Negative | 2-Negative | 2-Negative |
| 422 | 141 | 649 | Positive | Positive | Positive | 1-Positive | 1-Positive |
| 423 | 141 | 650 | Positive | Positive | Positive | 1-Positive | 1-Positive |
| 424 | 142 | 652 | Negative | Invalid | Negative | 3-Contami | 2-Negative |
| 425 | 142 | 653 | Negative | Negative | Negative | 2-Negative | 2-Negative |
| 426 | 142 | 651 | Negative | Negative | Negative | 2-Negative | 2-Negative |
| 427 | 143 | 656 | Positive | Negative | Negative | 3-Contami | 1-Positive |
| 428 | 143 | 654 | Positive | Positive | Positive | 2-Negative | 2-Negative |
| 429 | 143 | 655 | Positive | Negative | Negative | 3-Contami | 2-Negative |
| 430 | 144 | 661 | Negative | Negative | Negative | 2-Negative | 2-Negative |
| 431 | 144 | 659 | Negative | Positive | Negative | 3-Contami | 1-Positive |

| | | | | | | | |
|---|---|---|---|---|---|---|---|
| 432 | 144 | 660 | Negative | Negative | Negative | 3-Contami | 2-Negative |
| 433 | 145 | 657 | Negative | Negative | Negative | 2-Negative | 2-Negative |
| 434 | 145 | 658 | Negative | Negative | Negative | 2-Negative | 2-Negative |
| 435 | 145 | 662 | Negative | Positive | Negative | 2-Negative | 2-Negative |
| 436 | 146 | 644 | Positive | Positive | Positive | 1-Positive | 1-Positive |
| 437 | 146 | 642 | Positive | Negative | Negative | 2-Negative | 2-Negative |
| 438 | 146 | 633 | Positive | Invalid | Invalid | 2-Negative | 2-Negative |
| 439 | 147 | 641 | Negative | Negative | Negative | 2-Negative | 2-Negative |
| 440 | 147 | 643 | Negative | Negative | Negative | 2-Negative | 2-Negative |
| 441 | 147 | 629 | Negative | Negative | Negative | 2-Negative | 2-Negative |
| 442 | 148 | 630 | Negative | Negative | Negative | 2-Negative | 2-Negative |
| 443 | 148 | 631 | Negative | Negative | Negative | 3-Contami | 2-Negative |
| 444 | 148 | 634 | Negative | Negative | Negative | 2-Negative | 2-Negative |
| 445 | 149 | 636 | Positive | Negative | Negative | 2-Negative | 2-Negative |
| 446 | 149 | 637 | Positive | Positive | Positive | 2-Negative | 1-Positive |
| 447 | 149 | 635 | Positive | Negative | Negative | 2-Negative | 2-Negative |
| 448 | 150 | 702 | Negative | Negative | Negative | 2-Negative | 2-Negative |
| 449 | 150 | 703 | Negative | Negative | Negative | 3-Contami | 2-Negative |
| 450 | 150 | 704 | Negative | Positive | Negative | 2-Negative | 2-Negative |
| 451 | 151 | 705 | Negative | Negative | Negative | 2-Negative | 2-Negative |
| 452 | 151 | 708 | Negative | Negative | Negative | 2-Negative | 2-Negative |
| 453 | 151 | 701 | Negative | Negative | Negative | 2-Negative | 2-Negative |
| 454 | 152 | 707 | Negative | Negative | Negative | 2-Negative | 2-Negative |
| 455 | 152 | 709 | Negative | Negative | Negative | 2-Negative | 2-Negative |
| 456 | 152 | 712 | Negative | Negative | Negative | 2-Negative | 2-Negative |
| 457 | 153 | 714 | Negative | Positive | Negative | 2-Negative | 2-Negative |
| 458 | 153 | 713 | Negative | Negative | Negative | 3-Contami | 2-Negative |
| 459 | 153 | 706 | Negative | Negative | Negative | 2-Negative | 2-Negative |
| 460 | 154 | 719 | Negative | Negative | Negative | 2-Negative | 2-Negative |
| 461 | 154 | 711 | Negative | Negative | Negative | 2-Negative | 2-Negative |
| 462 | 154 | 710 | Negative | Negative | Negative | 2-Negative | 2-Negative |
| 463 | 155 | 718 | Negative | Negative | Negative | 2-Negative | 2-Negative |
| 464 | 155 | 717 | Negative | Positive | Negative | 2-Negative | 2-Negative |
| 465 | 155 | 715 | Negative | Negative | Negative | 2-Negative | 2-Negative |
| 466 | 156 | 722 | Negative | Negative | Negative | 2-Negative | 2-Negative |
| 467 | 156 | 723 | Negative | Negative | Negative | 2-Negative | 2-Negative |
| 468 | 156 | 716 | Negative | Negative | Negative | 2-Negative | 2-Negative |
| 469 | 157 | 724 | Positive | Positive | Positive | 2-Negative | 2-Negative |
| 470 | 157 | 721 | Positive | Negative | Negative | 2-Negative | 2-Negative |
| 471 | 157 | 725 | Positive | Positive | Positive | 1-Positive | 1-Positive |
| 472 | 158 | 247 | Negative | Negative | Negative | 2-Negative | 2-Negative |
| 473 | 158 | 720 | Negative | Negative | Negative | 2-Negative | 2-Negative |
| 474 | 158 | 248 | Negative | Negative | Negative | 2-Negative | 2-Negative |
| 475 | 159 | 244 | Positive | Negative | Negative | 2-Negative | 2-Negative |
| 476 | 159 | 118 | Positive | Positive | Positive | 1-Positive | 1-Positive |
| 477 | 159 | 245 | Positive | Positive | Positive | 1-Positive | 1-Positive |
| 478 | 160 | 663 | Negative | Negative | Negative | 2-Negative | 2-Negative |
| 479 | 160 | 249 | Negative | Negative | Negative | 2-Negative | 2-Negative |

| | | | | | | | |
|---|---|---|---|---|---|---|---|
| 480 | 160 | 482 | Negative | Negative | Negative | 2-Negative | 2-Negative |
| 481 | 161 | 250 | Negative | Positive | Negative | 2-Negative | 2-Negative |
| 482 | 161 | 249 | Negative | Negative | Negative | 2-Negative | 2-Negative |
| 483 | 161 | 113 | Negative | Negative | Negative | 2-Negative | 2-Negative |
| 484 | 162 | 251 | Negative | Negative | Negative | 3-Contami | 2-Negative |
| 485 | 162 | 121 | Negative | Negative | Negative | 2-Negative | 2-Negative |
| 486 | 162 | 112 | Negative | Negative | Negative | 2-Negative | 2-Negative |
| 487 | 163 | 639 | Negative | Negative | Negative | 2-Negative | 2-Negative |
| 488 | 163 | 640 | Negative | Negative | Negative | 2-Negative | 2-Negative |
| 489 | 163 | 632 | Negative | Negative | Negative | 3-Contami | 2-Negative |
| 490 | 164 | 402 | Negative | Negative | Negative | 2-Negative | 2-Negative |
| 491 | 164 | 401 | Negative | Negative | Negative | 2-Negative | 2-Negative |
| 492 | 164 | 638 | Negative | Negative | Negative | 2-Negative | 2-Negative |
| 493 | 165 | 405 | Positive | Positive | Positive | 1-Positive | 1-Positive |
| 494 | 165 | 404 | Positive | Negative | Negative | 2-Negative | 2-Negative |
| 495 | 165 | 403 | Positive | NoResult | NoResult | 2-Negative | 2-Negative |
| 496 | 166 | 494 | Negative | Negative | Negative | 2-Negative | 2-Negative |
| 497 | 166 | 406 | Negative | Negative | Negative | 2-Negative | 2-Negative |
| 498 | 166 | 407 | Negative | Negative | Negative | 2-Negative | 2-Negative |
| 499 | 167 | 283 | Negative | Negative | Negative | 3-Contami | 2-Negative |
| 500 | 167 | 285 | Negative | Positive | Negative | 1-Positive | 1-Positive |
| 501 | 167 | 284 | Negative | Negative | Negative | 2-Negative | 2-Negative |
| 502 | 168 | 492 | Negative | Negative | Negative | 2-Negative | 2-Negative |
| 503 | 168 | 495 | Negative | Negative | Negative | 2-Negative | 2-Negative |
| 504 | 168 | 493 | Negative | Negative | Negative | 2-Negative | 2-Negative |
| 505 | 169 | 271 | Positive | Positive | Positive | 1-Positive | 1-Positive |
| 506 | 169 | 272 | Positive | Negative | Negative | 2-Negative | 2-Negative |
| 507 | 169 | 273 | Positive | Negative | Negative | 2-Negative | 2-Negative |
| 508 | 170 | 276 | Negative | Negative | Negative | 2-Negative | 2-Negative |
| 509 | 170 | 275 | Negative | Negative | Negative | 2-Negative | 2-Negative |
| 510 | 170 | 274 | Negative | Negative | Negative | 2-Negative | 2-Negative |
| 511 | 171 | 488 | Negative | Negative | Negative | 2-Negative | 2-Negative |
| 512 | 171 | 489 | Negative | Negative | Negative | 2-Negative | 2-Negative |
| 513 | 171 | 490 | Negative | Negative | Negative | 2-Negative | 2-Negative |
| 514 | 172 | 278 | Negative | Negative | Negative | 2-Negative | 2-Negative |
| 515 | 172 | 131 | Negative | Negative | Negative | 2-Negative | 2-Negative |
| 516 | 172 | 277 | Negative | Negative | Negative | 2-Negative | 2-Negative |
| 517 | 173 | 279 | Positive | Negative | Negative | 1-Positive | 1-Positive |
| 518 | 173 | 664 | Positive | Negative | Negative | 2-Negative | 2-Negative |
| 519 | 173 | 665 | Positive | Negative | Negative | 3-Contami | 2-Negative |
| 520 | 174 | 666 | Negative | Positive | Negative | 2-Negative | 2-Negative |
| 521 | 174 | 668 | Negative | Negative | Negative | 2-Negative | 2-Negative |
| 522 | 174 | 667 | Negative | Negative | Negative | 2-Negative | 2-Negative |
| 523 | 175 | 132 | Negative | Negative | Negative | 2-Negative | 2-Negative |
| 524 | 175 | 670 | Negative | Invalid | Negative | 2-Negative | 2-Negative |
| 525 | 175 | 669 | Negative | Negative | Negative | 2-Negative | 2-Negative |
| 526 | 176 | 526 | Negative | Negative | Negative | 2-Negative | 2-Negative |
| 527 | 176 | 398 | Negative | Negative | Negative | 2-Negative | 2-Negative |

| | | | | | | | |
|---|---|---|---|---|---|---|---|
| 528 | 176 | 128 | Negative | Negative | Negative | 2-Negative | 2-Negative |
| 529 | 177 | 254 | Negative | Invalid | Negative | 2-Negative | 2-Negative |
| 530 | 177 | 252 | Negative | Negative | Negative | 3-Contami | 2-Negative |
| 531 | 177 | 255 | Negative | Negative | Negative | 2-Negative | 2-Negative |
| 532 | 178 | 124 | Negative | Negative | Negative | 2-Negative | 2-Negative |
| 533 | 178 | 256 | Negative | Negative | Negative | 2-Negative | 2-Negative |
| 534 | 178 | 397 | Negative | Negative | Negative | 2-Negative | 2-Negative |
| 535 | 179 | 125 | Negative | Negative | Negative | 2-Negative | 2-Negative |
| 536 | 179 | 122 | Negative | Negative | Negative | 2-Negative | 2-Negative |
| 537 | 179 | 258 | Negative | Negative | Negative | 2-Negative | 1-Positive |
| 538 | 180 | 260 | Negative | Negative | Negative | 2-Negative | 2-Negative |
| 539 | 180 | 261 | Negative | Negative | Negative | 2-Negative | 2-Negative |
| 540 | 180 | 259 | Negative | Negative | Negative | 2-Negative | 2-Negative |
| 541 | 181 | 523 | Negative | Negative | Negative | 2-Negative | 2-Negative |
| 542 | 181 | 127 | Negative | Negative | Negative | 2-Negative | 2-Negative |
| 543 | 181 | 483 | Negative | Negative | Negative | 2-Negative | 2-Negative |
| 544 | 182 | 395 | Negative | Negative | Negative | 2-Negative | 2-Negative |
| 545 | 182 | 522 | Negative | Negative | Negative | 2-Negative | 2-Negative |
| 546 | 182 | 257 | Negative | Negative | Negative | 2-Negative | 2-Negative |
| 547 | 183 | 269 | Negative | Negative | Negative | 3-Contami | 2-Negative |
| 548 | 183 | 267 | Negative | Negative | Negative | 2-Negative | 2-Negative |
| 549 | 183 | 268 | Negative | Negative | Negative | 2-Negative | 2-Negative |
| 550 | 184 | 524 | Negative | Negative | Negative | 2-Negative | 2-Negative |
| 551 | 184 | 270 | Negative | Negative | Negative | 2-Negative | 2-Negative |
| 552 | 184 | 525 | Negative | Negative | Negative | 3-Contami | 2-Negative |
| 553 | 185 | 396 | Negative | Negative | Negative | 2-Negative | 2-Negative |
| 554 | 185 | 486 | Negative | Negative | Negative | 2-Negative | 2-Negative |
| 555 | 185 | 485 | Negative | Negative | Negative | 3-Contami | 2-Negative |
| 556 | 186 | 130 | Invalid | Negative | Negative | 2-Negative | 2-Negative |
| 557 | 186 | 265 | Invalid | Positive | Positive | 2-Negative | 2-Negative |
| 558 | 186 | 487 | Invalid | Invalid | Invalid | 2-Negative | 2-Negative |
| 559 | 187 | 262 | Negative | Negative | Negative | 2-Negative | 2-Negative |
| 560 | 187 | 264 | Negative | Negative | Negative | 2-Negative | 2-Negative |
| 561 | 187 | 263 | Negative | Negative | Negative | 2-Negative | 2-Negative |
| 562 | 188 | 280 | Negative | Negative | Negative | 2-Negative | 2-Negative |
| 563 | 188 | 266 | Negative | Negative | Negative | 2-Negative | 2-Negative |
| 564 | 188 | 281 | Negative | Negative | Negative | 2-Negative | 2-Negative |
| 565 | 189 | 399 | Positive | Positive | Positive | 1-Positive | 1-Positive |
| 566 | 189 | 400 | Positive | Negative | Negative | 3-Contami | 2-Negative |
| 567 | 189 | 527 | Positive | Positive | Positive | 1-Positive | 1-Positive |
| 568 | 190 | 133 | Positive | Negative | Negative | 2-Negative | 2-Negative |
| 569 | 190 | 220 | Positive | Negative | Negative | 2-Negative | 2-Negative |
| 570 | 190 | 282 | Positive | Positive | Positive | 1-Positive | 1-Positive |
| 571 | 191 | 221 | Positive | Positive | Positive | 1-Positive | 1-Positive |
| 572 | 191 | 115 | Positive | Negative | Negative | 2-Negative | 2-Negative |
| 573 | 191 | 111 | Positive | Negative | Negative | 2-Negative | 2-Negative |
| 574 | 192 | 116 | Negative | Negative | Negative | 2-Negative | 2-Negative |
| 575 | 192 | 117 | Negative | Negative | Negative | 2-Negative | 2-Negative |

| | | | | | | | |
|---|---|---|---|---|---|---|---|
| 576 | 192 | 246 | Negative | Negative | Negative | 2-Negative | 2-Negative |
| 577 | 193 | 119 | Negative | Negative | Negative | 2-Negative | 2-Negative |
| 578 | 193 | 114 | Negative | Negative | Negative | 2-Negative | 2-Negative |
| 579 | 193 | 120 | Negative | NoResult | Negative | 2-Negative | 2-Negative |
| 580 | 194 | 812 | Negative | Invalid | Negative | 2-Negative | 2-Negative |
| 581 | 194 | 814 | Negative | Negative | Negative | 2-Negative | 2-Negative |
| 582 | 194 | 811 | Negative | Negative | Negative | 2-Negative | 2-Negative |
| 583 | 195 | 810 | Negative | Negative | Negative | 2-Negative | 2-Negative |
| 584 | 195 | 813 | Negative | Negative | Negative | 2-Negative | 2-Negative |
| 585 | 195 | 823 | Negative | Negative | Negative | 2-Negative | 2-Negative |
| 586 | 196 | 815 | Negative | Negative | Negative | 2-Negative | 2-Negative |
| 587 | 196 | 822 | Negative | Negative | Negative | 2-Negative | 2-Negative |
| 588 | 196 | 824 | Negative | Negative | Negative | 2-Negative | 2-Negative |
| 589 | 197 | 819 | Negative | Negative | Negative | 2-Negative | 2-Negative |
| 590 | 197 | 818 | Negative | Negative | Negative | 2-Negative | 2-Negative |
| 591 | 197 | 817 | Negative | Negative | Negative | 2-Negative | 2-Negative |
| 592 | 198 | 821 | Negative | Negative | Negative | 2-Negative | 2-Negative |
| 593 | 198 | 816 | Negative | Negative | Negative | 3-Contami | 2-Negative |
| 594 | 198 | 820 | Negative | Negative | Negative | 2-Negative | 2-Negative |
| 595 | 199 | 327 | Negative | Negative | Negative | 2-Negative | 2-Negative |
| 596 | 199 | 326 | Negative | Negative | Negative | 2-Negative | 2-Negative |
| 597 | 199 | 328 | Negative | Positive | Negative | 2-Negative | 1-Positive |
| 598 | 200 | 322 | Negative | Negative | Negative | 2-Negative | 2-Negative |
| 599 | 200 | 324 | Negative | Negative | Negative | 2-Negative | 2-Negative |
| 600 | 200 | 323 | Negative | Negative | Negative | 2-Negative | 2-Negative |
| 601 | 201 | 547 | Negative | Negative | Negative | 2-Negative | 2-Negative |
| 602 | 201 | 545 | Negative | Negative | Negative | 2-Negative | 2-Negative |
| 603 | 201 | 546 | Negative | Negative | Negative | 2-Negative | 2-Negative |
| 604 | 202 | 331 | Negative | Negative | Negative | 2-Negative | 2-Negative |
| 605 | 202 | 330 | Negative | Negative | Negative | 2-Negative | 2-Negative |
| 606 | 202 | 333 | Negative | Negative | Negative | 2-Negative | 2-Negative |
| 607 | 203 | 845 | Negative | Negative | Negative | 2-Negative | 2-Negative |
| 608 | 203 | 329 | Negative | Negative | Negative | 2-Negative | 1-Positive |
| 609 | 203 | 332 | Negative | Negative | Negative | 2-Negative | 2-Negative |
| 610 | 204 | 846 | Negative | Negative | Negative | 2-Negative | 2-Negative |
| 611 | 204 | 843 | Negative | Negative | Negative | 2-Negative | 2-Negative |
| 612 | 204 | 844 | Negative | Negative | Negative | 3-Contami | 2-Negative |
| 613 | 205 | 848 | Negative | Negative | Negative | 2-Negative | 2-Negative |
| 614 | 205 | 849 | Negative | Negative | Negative | 3-Contami | 2-Negative |
| 615 | 205 | 847 | Negative | Negative | Negative | 2-Negative | 2-Negative |
| 616 | 206 | 837 | Negative | Negative | Negative | 2-Negative | 2-Negative |
| 617 | 206 | 836 | Negative | Negative | Negative | 2-Negative | 2-Negative |
| 618 | 206 | 838 | Negative | Negative | Negative | 2-Negative | 2-Negative |
| 619 | 207 | 840 | Negative | Negative | Negative | 2-Negative | 2-Negative |
| 620 | 207 | 839 | Negative | Negative | Negative | 2-Negative | 2-Negative |
| 621 | 207 | 841 | Negative | Negative | Negative | 2-Negative | 1-Positive |
| 622 | 208 | 835 | Negative | Negative | Negative | 2-Negative | 2-Negative |
| 623 | 208 | 834 | Negative | Positive | Negative | 2-Negative | 2-Negative |

| | | | | | | | |
|---|---|---|---|---|---|---|---|
| 624 | 208 | 842 | Negative | Negative | Negative | 2-Negative | 2-Negative |
| 625 | 209 | 833 | Negative | Negative | Negative | 2-Negative | 2-Negative |
| 626 | 209 | 549 | Negative | Negative | Negative | 2-Negative | 2-Negative |
| 627 | 209 | 550 | Negative | Negative | Negative | 2-Negative | 2-Negative |
| 628 | 210 | 502 | Negative | Negative | Negative | 2-Negative | 2-Negative |
| 629 | 210 | 548 | Negative | Negative | Negative | 2-Negative | 2-Negative |
| 630 | 210 | 551 | Negative | Negative | Negative | 2-Negative | 2-Negative |
| 631 | 211 | 507 | Negative | Negative | Negative | 2-Negative | 2-Negative |
| 632 | 211 | 411 | Negative | Negative | Negative | 2-Negative | 2-Negative |
| 633 | 211 | 506 | Negative | Negative | Negative | 2-Negative | 2-Negative |
| 634 | 212 | 851 | Negative | Negative | Negative | 2-Negative | 2-Negative |
| 635 | 212 | 804 | Negative | Negative | Negative | 2-Negative | 2-Negative |
| 636 | 212 | 852 | Negative | Negative | Negative | 2-Negative | 2-Negative |
| 637 | 213 | 854 | Negative | Negative | Negative | 2-Negative | 2-Negative |
| 638 | 213 | 853 | Negative | Positive | Negative | 2-Negative | 2-Negative |
| 639 | 213 | 850 | Negative | Negative | Negative | 2-Negative | 2-Negative |
| 640 | 214 | 828 | Positive | Negative | Negative | 2-Negative | 2-Negative |
| 641 | 214 | 855 | Positive | Negative | Negative | 2-Negative | 2-Negative |
| 642 | 214 | 829 | Positive | Positive | Positive | 1-Positive | 1-Positive |
| 643 | 215 | 831 | Negative | Negative | Negative | 2-Negative | 2-Negative |
| 644 | 215 | 825 | Negative | Negative | Negative | 2-Negative | 2-Negative |
| 645 | 215 | 826 | Negative | Negative | Negative | 2-Negative | 2-Negative |
| 646 | 216 | 505 | Negative | Negative | Negative | 2-Negative | 2-Negative |
| 647 | 216 | 827 | Negative | Negative | Negative | 2-Negative | 2-Negative |
| 648 | 216 | 153 | Negative | Negative | Negative | 2-Negative | 2-Negative |
| 649 | 217 | 316 | Negative | Negative | Negative | 2-Negative | 2-Negative |
| 650 | 217 | 543 | Negative | Negative | Negative | 2-Negative | 1-Positive |
| 651 | 217 | 544 | Negative | Negative | Negative | 2-Negative | 2-Negative |
| 652 | 218 | 317 | Negative | Negative | Negative | 2-Negative | 2-Negative |
| 653 | 218 | 318 | Negative | Negative | Negative | 2-Negative | 2-Negative |
| 654 | 218 | 319 | Negative | Negative | Negative | 2-Negative | 2-Negative |
| 655 | 219 | 320 | Negative | Negative | Negative | 2-Negative | 2-Negative |
| 656 | 219 | 321 | Negative | Positive | Negative | 2-Negative | 2-Negative |
| 657 | 219 | 539 | Negative | Negative | Negative | 3-Contami | 2-Negative |
| 658 | 220 | 317 | Negative | Negative | Negative | 2-Negative | 2-Negative |
| 659 | 220 | 806 | Negative | Negative | Negative | 2-Negative | 2-Negative |
| 660 | 220 | 807 | Negative | Invalid | Negative | 2-Negative | 2-Negative |
| 661 | 221 | 745 | Positive | Negative | Negative | 2-Negative | 1-Positive |
| 662 | 221 | 746 | Positive | Negative | Negative | 2-Negative | 2-Negative |
| 663 | 221 | 503 | Positive | Negative | Negative | 2-Negative | 2-Negative |
| 664 | 222 | 747 | Negative | Negative | Negative | 2-Negative | 2-Negative |
| 665 | 222 | 748 | Negative | Negative | Negative | 2-Negative | 2-Negative |
| 666 | 222 | 759 | Negative | Negative | Negative | 2-Negative | 2-Negative |
| 667 | 223 | 750 | Negative | Negative | Negative | 2-Negative | 2-Negative |
| 668 | 223 | 749 | Negative | Negative | Negative | 2-Negative | 2-Negative |
| 669 | 223 | 751 | Negative | Negative | Negative | 2-Negative | 2-Negative |
| 670 | 224 | 756 | Negative | Negative | Negative | 2-Negative | 2-Negative |
| 671 | 224 | 757 | Negative | Negative | Negative | 2-Negative | 2-Negative |

| | | | | | | | |
|---|---|---|---|---|---|---|---|
| 672 | 224 | 755 | Negative | Negative | Negative | 1-Positive | 2-Negative |
| 673 | 225 | 754 | Positive | Negative | Negative | 2-Negative | 2-Negative |
| 674 | 225 | 753 | Positive | Positive | Positive | 2-Negative | 1-Positive |
| 675 | 225 | 752 | Positive | Negative | Negative | 2-Negative | 2-Negative |
| 676 | 226 | 762 | Negative | Negative | Negative | 3-Contami | 2-Negative |
| 677 | 226 | 761 | Negative | Negative | Negative | 3-Contami | 1-Positive |
| 678 | 226 | 763 | Negative | Negative | Negative | 2-Negative | 2-Negative |
| 679 | 227 | 759 | Negative | Negative | Negative | 2-Negative | 2-Negative |
| 680 | 227 | 760 | Negative | Negative | Negative | 2-Negative | 2-Negative |
| 681 | 227 | 758 | Negative | Negative | Negative | 2-Negative | 2-Negative |
| 682 | 228 | 764 | Negative | Negative | Negative | 2-Negative | 2-Negative |
| 683 | 228 | 765 | Negative | Negative | Negative | 2-Negative | 2-Negative |
| 684 | 228 | 768 | Negative | Negative | Negative | 2-Negative | 2-Negative |
| 685 | 229 | 767 | Positive | Negative | Negative | 2-Negative | 2-Negative |
| 686 | 229 | 766 | Positive | Positive | Positive | 1-Positive | 1-Positive |
| 687 | 229 | 769 | Positive | Negative | Negative | 2-Negative | 2-Negative |
| 688 | 230 | 771 | Positive | Positive | Positive | 1-Positive | 1-Positive |
| 689 | 230 | 294 | Positive | Positive | Positive | 1-Positive | 1-Positive |
| 690 | 230 | 770 | Positive | Negative | Negative | 2-Negative | 2-Negative |
| 691 | 231 | 532 | Positive | Negative | Negative | 2-Negative | 2-Negative |
| 692 | 231 | 533 | Positive | Negative | Negative | 3-Contami | 2-Negative |
| 693 | 231 | 295 | Positive | Positive | Positive | 1-Positive | 1-Positive |
| 694 | 232 | 409 | Negative | Negative | Negative | 2-Negative | 2-Negative |
| 695 | 232 | 830 | Negative | Negative | Negative | 2-Negative | 2-Negative |
| 696 | 232 | 146 | Negative | Negative | Negative | 2-Negative | 2-Negative |
| 697 | 233 | 832 | Positive | Negative | Negative | 2-Negative | 2-Negative |
| 698 | 233 | 734 | Positive | Negative | Negative | 2-Negative | 2-Negative |
| 699 | 233 | 529 | Positive | Positive | Positive | 1-Positive | 1-Positive |
| 700 | 234 | 736 | Positive | Positive | Positive | 1-Positive | 1-Positive |
| 701 | 234 | 738 | Positive | Negative | Negative | 2-Negative | 2-Negative |
| 702 | 234 | 735 | Positive | Negative | Negative | 2-Negative | 2-Negative |
| 703 | 235 | 739 | Negative | Negative | Negative | 2-Negative | 2-Negative |
| 704 | 235 | 740 | Negative | Invalid | Negative | 2-Negative | 2-Negative |
| 705 | 235 | 737 | Negative | Negative | Negative | 2-Negative | 2-Negative |
| 706 | 236 | 742 | Negative | Negative | Negative | 2-Negative | 2-Negative |
| 707 | 236 | 129 | Negative | Negative | Negative | 2-Negative | 2-Negative |
| 708 | 236 | 741 | Negative | Negative | Negative | 2-Negative | 2-Negative |
| 709 | 237 | 528 | Negative | Negative | Negative | 2-Negative | 2-Negative |
| 710 | 237 | 730 | Negative | Negative | Negative | 2-Negative | 2-Negative |
| 711 | 237 | 134 | Negative | Negative | Negative | 2-Negative | 2-Negative |
| 712 | 238 | 729 | Negative | Negative | Negative | 2-Negative | 2-Negative |
| 713 | 238 | 727 | Negative | Negative | Negative | 2-Negative | 2-Negative |
| 714 | 238 | 728 | Negative | Negative | Negative | 2-Negative | 2-Negative |
| 715 | 239 | 732 | Positive | Negative | Negative | 2-Negative | 2-Negative |
| 716 | 239 | 731 | Positive | Positive | Positive | 1-Positive | 1-Positive |
| 717 | 239 | 726 | Positive | Positive | Positive | 1-Positive | 1-Positive |
| 718 | 240 | 733 | Positive | Negative | Negative | 2-Negative | 2-Negative |
| 719 | 240 | 496 | Positive | Positive | Positive | 1-Positive | 1-Positive |

| | | | | | | | |
|---|---|---|---|---|---|---|---|
| 720 | 240 | 135 | Positive | Negative | Negative | 2-Negative | 2-Negative |
| 721 | 241 | 530 | Positive | Negative | Negative | 2-Negative | 2-Negative |
| 722 | 241 | 136 | Positive | Positive | Positive | 1-Positive | 1-Positive |
| 723 | 241 | 497 | Positive | Negative | Negative | 2-Negative | 2-Negative |
| 724 | 242 | 288 | Positive | Negative | Negative | 2-Negative | 2-Negative |
| 725 | 242 | 531 | Positive | Positive | Positive | 1-Positive | 1-Positive |
| 726 | 242 | 808 | Positive | Negative | Negative | 2-Negative | 2-Negative |
| 727 | 243 | 415 | Negative | Negative | Negative | 2-Negative | 2-Negative |
| 728 | 243 | 414 | Negative | Negative | Negative | 2-Negative | 2-Negative |
| 729 | 243 | 809 | Negative | Negative | Negative | 2-Negative | 2-Negative |
| 730 | 244 | 315 | Positive | Positive | Positive | 1-Positive | 1-Positive |
| 731 | 244 | 314 | Positive | Positive | Positive | 1-Positive | 1-Positive |
| 732 | 244 | 791 | Positive | Negative | Negative | 2-Negative | 2-Negative |
| 733 | 245 | 790 | Negative | Negative | Negative | 2-Negative | 2-Negative |
| 734 | 245 | 792 | Negative | Negative | Negative | 2-Negative | 2-Negative |
| 735 | 245 | 789 | Negative | Negative | Negative | 2-Negative | 2-Negative |
| 736 | 246 | 795 | Positive | Negative | Negative | 2-Negative | 2-Negative |
| 737 | 246 | 534 | Positive | Negative | Negative | 2-Negative | 2-Negative |
| 738 | 246 | 535 | Positive | Negative | Negative | 2-Negative | 2-Negative |
| 739 | 247 | 538 | Negative | Negative | Negative | 2-Negative | 2-Negative |
| 740 | 247 | 536 | Negative | Negative | Negative | 2-Negative | 2-Negative |
| 741 | 247 | 537 | Negative | Negative | Negative | 2-Negative | 2-Negative |
| 742 | 248 | 793 | Negative | Negative | Negative | 2-Negative | 2-Negative |
| 743 | 248 | 781 | Negative | Negative | Negative | 3-Contami | 2-Negative |
| 744 | 248 | 782 | Negative | Negative | Negative | 2-Negative | 2-Negative |
| 745 | 249 | 787 | Positive | Negative | Negative | 9-Missing | 1-Positive |
| 746 | 249 | 785 | Positive | Negative | Negative | 2-Negative | 2-Negative |
| 747 | 249 | 783 | Positive | Positive | Positive | 1-Positive | 1-Positive |
| 748 | 250 | 786 | Negative | Negative | Negative | 2-Negative | 2-Negative |
| 749 | 250 | 784 | Negative | Negative | Negative | 2-Negative | 2-Negative |
| 750 | 250 | 788 | Negative | Negative | Negative | 2-Negative | 2-Negative |
| 751 | 251 | 287 | Positive | Positive | Positive | 1-Positive | 1-Positive |
| 752 | 251 | 289 | Positive | Positive | Positive | 1-Positive | 1-Positive |
| 753 | 251 | 286 | Positive | Positive | Positive | 1-Positive | 1-Positive |
| 754 | 252 | 291 | Negative | Negative | Negative | 2-Negative | 2-Negative |
| 755 | 252 | 290 | Negative | Negative | Negative | 2-Negative | 2-Negative |
| 756 | 252 | 292 | Negative | Negative | Negative | 2-Negative | 2-Negative |
| 757 | 253 | 501 | Negative | Negative | Negative | 2-Negative | 2-Negative |
| 758 | 253 | 293 | Negative | Negative | Negative | 3-Contami | 2-Negative |
| 759 | 253 | 498 | Negative | Negative | Negative | 2-Negative | 2-Negative |
| 760 | 254 | 296 | Negative | Negative | Negative | 2-Negative | 2-Negative |
| 761 | 254 | 145 | Negative | Negative | Negative | 2-Negative | 2-Negative |
| 762 | 254 | 144 | Negative | Negative | Negative | 2-Negative | 2-Negative |
| 763 | 255 | 300 | Positive | Positive | Positive | 1-Positive | 1-Positive |
| 764 | 255 | 301 | Positive | Positive | Positive | 1-Positive | 1-Positive |
| 765 | 255 | 297 | Positive | Negative | Negative | 2-Negative | 2-Negative |
| 766 | 256 | 305 | Negative | Negative | Negative | 2-Negative | 2-Negative |
| 767 | 256 | 410 | Negative | Negative | Negative | 2-Negative | 2-Negative |

| | | | | | | | |
|---|---|---|---|---|---|---|---|
| 768 | 256 | 298 | Negative | Negative | Negative | 2-Negative | 1-Positive |
| 769 | 257 | 304 | Negative | Negative | Negative | 2-Negative | 2-Negative |
| 770 | 257 | 149 | Negative | Negative | Negative | 2-Negative | 2-Negative |
| 771 | 257 | 148 | Negative | Negative | Negative | 3-Contami | 2-Negative |
| 772 | 258 | 541 | Positive | Positive | Positive | 1-Positive | 1-Positive |
| 773 | 258 | 540 | Positive | Negative | Negative | 2-Negative | 2-Negative |
| 774 | 258 | 542 | Positive | Negative | Negative | 2-Negative | 2-Negative |
| 775 | 259 | 412 | Negative | Negative | Negative | 2-Negative | 2-Negative |
| 776 | 259 | 413 | Negative | Negative | Negative | 2-Negative | 2-Negative |
| 777 | 259 | 308 | Negative | Negative | Negative | 2-Negative | 2-Negative |
| 778 | 260 | 307 | Negative | Negative | Negative | 2-Negative | 2-Negative |
| 779 | 260 | 309 | Negative | Negative | Negative | 2-Negative | 2-Negative |
| 780 | 260 | 310 | Negative | Negative | Negative | 2-Negative | 2-Negative |
| 781 | 261 | 802 | Negative | Negative | Negative | 2-Negative | 2-Negative |
| 782 | 261 | 800 | Negative | Negative | Negative | 2-Negative | 2-Negative |
| 783 | 261 | 803 | Negative | Negative | Negative | 2-Negative | 2-Negative |
| 784 | 262 | 799 | Positive | Negative | Negative | 2-Negative | 2-Negative |
| 785 | 262 | 798 | Positive | Positive | Positive | 1-Positive | 1-Positive |
| 786 | 262 | 801 | Positive | Negative | Negative | 3-Contami | 2-Negative |
| 787 | 263 | 797 | Negative | Negative | Negative | 3-Contami | 2-Negative |
| 788 | 263 | 306 | Negative | Negative | Negative | 2-Negative | 2-Negative |
| 789 | 263 | 796 | Negative | Negative | Negative | 2-Negative | 2-Negative |
| 790 | 264 | 780 | Positive | Positive | Positive | 3-Contami | 2-Negative |
| 791 | 264 | 147 | Positive | Negative | Negative | 2-Negative | 2-Negative |
| 792 | 264 | 779 | Positive | Invalid | Invalid | 2-Negative | 2-Negative |
| 793 | 265 | 152 | Negative | Negative | Negative | 2-Negative | 2-Negative |
| 794 | 265 | 312 | Negative | Negative | Negative | 2-Negative | 2-Negative |
| 795 | 265 | 778 | Negative | Negative | Negative | 3-Contami | 2-Negative |
| 796 | 266 | 140 | Positive | Negative | Negative | 2-Negative | 2-Negative |
| 797 | 266 | 150 | Positive | Negative | Negative | 2-Negative | 2-Negative |
| 798 | 266 | 151 | Positive | Positive | Positive | 2-Negative | 2-Negative |
| 799 | 267 | 141 | Positive | Negative | Negative | 3-Contami | 2-Negative |
| 800 | 267 | 311 | Positive | Positive | Positive | 1-Positive | 1-Positive |
| 801 | 267 | 794 | Positive | Negative | Negative | 2-Negative | 2-Negative |
| 802 | 268 | 64 | Negative | Negative | Negative | 2-Negative | 2-Negative |
| 803 | 268 | 390 | Negative | Negative | Negative | 2-Negative | 2-Negative |
| 804 | 268 | 60 | Negative | Negative | Negative | 2-Negative | 2-Negative |
| 805 | 269 | 242 | Positive | Negative | Negative | 2-Negative | 2-Negative |
| 806 | 269 | 384 | Positive | Negative | Negative | 2-Negative | 2-Negative |
| 807 | 269 | 463 | Positive | Negative | Negative | 2-Negative | 2-Negative |
| 808 | 270 | 96 | Positive | Negative | Negative | 2-Negative | 2-Negative |
| 809 | 270 | 108 | Positive | Positive | Positive | 1-Positive | 1-Positive |
| 810 | 270 | 225 | Positive | Negative | Negative | 2-Negative | 2-Negative |
| 811 | 271 | 419 | Positive | Positive | Positive | 1-Positive | 1-Positive |
| 812 | 271 | 509 | Positive | Positive | Positive | 1-Positive | 1-Positive |
| 813 | 271 | 510 | Positive | Negative | Negative | 2-Negative | 2-Negative |
| 814 | 272 | 418 | Positive | Negative | Negative | 2-Negative | 2-Negative |
| 815 | 272 | 508 | Positive | Positive | Positive | 1-Positive | 1-Positive |

| | | | | | | | |
|---|---|---|---|---|---|---|---|
| 816 | 272 | 154 | Positive | Negative | Negative | 3-Contami | 2-Negative |
| 817 | 273 | 416 | Negative | Negative | Negative | 3-Contami | 2-Negative |
| 818 | 273 | 417 | Negative | Negative | Negative | 2-Negative | 2-Negative |
| 819 | 273 | 500 | Negative | Negative | Negative | 2-Negative | 2-Negative |
| 820 | 274 | 774 | Negative | Negative | Negative | 2-Negative | 2-Negative |
| 821 | 274 | 773 | Negative | Negative | Negative | 2-Negative | 2-Negative |
| 822 | 274 | 499 | Negative | Negative | Negative | 3-Contami | 2-Negative |
| 823 | 275 | 553 | Negative | Negative | Negative | 3-Contami | 2-Negative |
| 824 | 275 | 772 | Negative | Negative | Negative | 2-Negative | 2-Negative |
| 825 | 275 | 552 | Negative | Negative | Negative | 2-Negative | 2-Negative |
| 826 | 276 | 313 | Negative | Negative | Negative | 2-Negative | 2-Negative |
| 827 | 276 | 302 | Negative | Negative | Negative | 2-Negative | 2-Negative |
| 828 | 276 | 325 | Negative | Negative | Negative | 2-Negative | 2-Negative |
| 829 | 277 | 504 | Negative | Negative | Negative | 2-Negative | 2-Negative |
| 830 | 277 | 743 | Negative | Negative | Negative | 2-Negative | 2-Negative |
| 831 | 277 | 744 | Negative | Negative | Negative | 2-Negative | 2-Negative |
| 832 | 278 | 776 | Negative | Negative | Negative | 2-Negative | 2-Negative |
| 833 | 278 | 775 | Negative | Negative | Negative | 2-Negative | 2-Negative |
| 834 | 278 | 777 | Negative | Invalid | Negative | 2-Negative | 2-Negative |
| 835 | 279 | 137 | Negative | Negative | Negative | 2-Negative | 2-Negative |
| 836 | 279 | 143 | Negative | Negative | Negative | 2-Negative | 2-Negative |
| 837 | 279 | 142 | Negative | Negative | Negative | 2-Negative | 2-Negative |
| 838 | 280 | 408 | Positive | Positive | Positive | 1-Positive | 1-Positive |
| 839 | 280 | 139 | Positive | Negative | Negative | 2-Negative | 2-Negative |
| 840 | 280 | 299 | Positive | Negative | Negative | 2-Negative | 2-Negative |
| 841 | 281 | 491 | Positive | Negative | Negative | 2-Negative | 2-Negative |
| 842 | 281 | 580 | Positive | Negative | Negative | 2-Negative | 2-Negative |
| 843 | 281 | 381 | Positive | NoResult | NoResult | 1-Positive | 1-Positive |
| 844 | 282 | 461 | Negative | Negative | Negative | 2-Negative | 2-Negative |
| 845 | 282 | 126 | Negative | Negative | Negative | 2-Negative | 2-Negative |
| 846 | 282 | 123 | Negative | Negative | Negative | 2-Negative | 2-Negative |
| 847 | 283 | 253 | Positive | Negative | Negative | 1-Positive | 1-Positive |
| 848 | 283 | 585 | Positive | Invalid | Invalid | 3-Contami | 1-Positive |
| 849 | 283 | 138 | Positive | Negative | Negative | 2-Negative | 2-Negative |
| 850 | 284 | 160 | Invalid | Negative | Negative | 2-Negative | 2-Negative |
| 851 | 284 | 337 | Invalid | Negative | Negative | 2-Negative | 2-Negative |
| 852 | 284 | 342 | Invalid | Negative | Negative | 2-Negative | 2-Negative |
| 853 | 285 | 189 | Positive | Positive | Positive | 1-Positive | 1-Positive |
| 854 | 285 | 190 | Positive | Negative | Negative | 1-Positive | 2-Negative |
| 855 | 285 | 171 | Positive | Positive | Positive | 2-Negative | 2-Negative |
| 856 | 286 | 359 | Positive | Positive | Positive | 1-Positive | 1-Positive |
| 857 | 286 | 3 | Positive | Positive | Positive | 1-Positive | 1-Positive |
| 858 | 286 | 357 | Positive | Positive | Positive | 1-Positive | 1-Positive |
| 859 | 287 | 303 | Negative | Negative | Negative | 2-Negative | 2-Negative |
| 860 | 287 | 805 | Negative | Negative | Negative | 2-Negative | 2-Negative |
| 861 | 287 | 91 | Negative | Negative | Negative | 2-Negative | 2-Negative |
